# Selection of Genetic Conditions for Multi-State Genomic Newborn Screening in BEACONS-NBS

**DOI:** 10.64898/2026.03.23.26349079

**Authors:** Nina B. Gold, Britt A. Johnson, Harini Somanchi, Thomas Minten, Stephanie A. Coury, Carrie Blout Zawatsky, Amber Begtrup, Elizabeth Butler, Katherine G. Langley, Rebekah Zimmerman, Heather M. McLaughlin, Tessa Ellefson, Adriann Kern, Heidi L. Rehm, David Bick, Steven E. Brenner, Dalia Kasperaviciute, Roshini S. Abraham, Ivona Aksentijevich, Melanie Babinski, Charles J. Billington, Manish J. Butte, Scott W. Canna, Makena Caron, Yee-Ming Chan, Shanmuganathan Chandrakasan, Samuel C.C. Chiang, Ottavia M. Delmonte, Lisa R. Diller, Lilian Downie, Julie Fleischer, Anne Fulton, Rebecca D. Ganetzky, Jessica Gold, Raphaela Goldbach-Mansky, Eyal Grunebaum, Rebecca C. Hale, Ada Hamosh, Friedhelm Hildebrandt, Alexander M. Holtz, Christina Jacobsen, Matthew J. Kan, Maleewan Kitcharoensakkul, Vilmante Kodyte, Raymond J. Kreienkamp, Belinda S. Lennerz, Angela E. Lin, Adithya V. Madduri, Joseph A. Majzoub, Rebecca A. Marsh, David T. Miller, Joshua D. Milner, Deborah M. Mitchell, Brittney Murray, Luigi D. Notarangelo, Xiao P. Peng, Emily M. Place, Craig D. Platt, Sharon E. Plon, Scott R. Plotkin, Cynthia M. Powell, Jennifer M. Puck, Amy E. Roberts, Vijay G. Sankaran, Daniella M. Schwartz, Christine M. Seroogy, Shreyas Srinath, Alanna Strong, Kathleen E. Sullivan, Weizhen Tan, Teresa K. Tarrant, Jay R. Thiagarajah, Melissa A. Walker, Ari J. Wassner, Hannie Zomer Bolanos, The BEACONS-NBS team, Amy M. Gaviglio, Natasha Bonhomme, Jelili Ojodu, Sikha Singh, Guisou Zarbalian, Stacey Pereira, Aaron J. Goldenberg, Ingrid A. Holm, Melissa P. Wasserstein, Robert C. Green

## Abstract

**Introduction:** BEACONS-NBS (Building Evidence and Collaboration for GenOmics in Nationwide Newborn Screening) is the first research study to integrate whole genome sequencing into newborn screening (NBS) across multiple U.S. states and territorial public health laboratory programs (PHLPs). We developed a list of conditions for screening.

**Methods:** We designed inclusion criteria and assembled an initial condition list from published resources. The list was revised by clinical experts, molecular geneticists, genetic counselors, PHLPs, rare disease advocacy organizations, the BEACONS-NBS Community Advisory Board, and project leadership from the National Institutes of Health. For each condition, we provided a rationale for early detection, diagnostic signs or biomarkers, and treatments or surveillance strategies.

**Results:** The BEACONS-NBS condition list includes 777 conditions associated with 743 genes, one copy number variant, and two aneuploidies and is larger than those used in other genomic NBS research studies in the U.S. and United Kingdom. Most conditions are inborn errors of immunity (37.2%), inherited metabolic disorders (18.7%), or endocrine conditions (18.1%). Nearly all conditions (93.3%) can be confirmed using a non-genetic test.

**Discussion:** BEACONS-NBS has established a condition list for implementation across multiple state and territorial PHLPs, enabling the prospective evaluation of feasibility of population-wide genomic NBS.

## Introduction

Newborn screening (NBS) is a public health program that detects approximately 15,000 infants at risk for treatable conditions in the United States each year.^1,2^ The addition of conditions to NBS has been largely guided by the historic Wilson-Jungner principles for population screening, which emphasize the inclusion of treatable, childhood-onset conditions.^3^ Most conditions included in NBS are identified using biochemical laboratory markers measurable in dried blood spots. However, many other conditions that are potentially eligible for population screening lack suitable biochemical markers and therefore require alternative screening approaches. Genetic information is increasingly used as both a first- and second-tier screening method for select disorders,^4,5^ but only a fraction of the thousands of known genetic conditions are currently included in state NBS programs.^6,7^

Over 30 international research programs are exploring genomic testing, including whole genome sequencing (WGS), as a strategy to identify newborns with additional conditions.^8,9^ Although WGS expands the diagnostic potential of NBS, determining which conditions should be included is ethically and scientifically complex. Considerations include the strength of the gene-condition association,^10^ penetrance, prevalence, age of onset, severity, availability of a confirmatory non-genetic diagnostic test, and efficacy of treatment or surveillance.^11^ Although some of these characteristics have been quantified,^12,13^ others remain subjective, contributing to the heterogeneity of conditions included in genomic NBS research programs.^9,14^ Surveys of genetics professionals and the public,^15,16^ qualitative studies involving parents,^17–19^ analyses of genomic NBS research studies,^9,20,21^ and recommendations from the International Consortium of Newborn Sequencing (ICoNS)^22^ have shaped general selection principles and proposed specific high-priority conditions for genomic NBS; however, specific consensus guidelines are lacking.

In March 2025, the National Institutes of Health (NIH) Common Fund Venture Initiative issued an Other Transaction Authority (OTA) research opportunity announcement for the Newborn Screening by Whole Genome Sequencing (NBSxWGS) Collaboratory, aimed at evaluating the feasibility of using WGS as a first-tier screening test for selected genetic conditions actionable in the first year of life. Our group received this award and launched BEACONS-NBS (Building Evidence and Collaboration for GenOmics in Nationwide Newborn Screening), a three-year study assessing integration of WGS into NBS across public health laboratory programs (PHLPs) in seven U.S. states and territories: Iowa, Minnesota, New York, Oregon, Puerto Rico, South Carolina, and Texas.

Here we describe the process of developing a list of genetic conditions to be analyzed in up to 30,000 infants enrolled in BEACONS-NBS study.

## Methods

### Inclusion criteria for the BEACONS-NBS condition list

<u>BEACONS-NBS condition list committee.</u> The BEACONS-NBS condition list committee was led by a medical biochemical geneticist who is one of the BEACONS-NBS four principal investigators (N.B.G.) and a clinical biochemical and molecular geneticist (B.A.J.), with program management from a genetic counselor (S.A.C.) and a clinical research coordinator (H.S.). The committee also included another BEACONS-NBS principal investigator (I.A.H.), clinical molecular geneticists and genetic counselors from GeneDx (A.B., K.G.L., R.Z.), consultants with expertise in molecular genetics and genomic NBS (H.L.R., D.B., S.E.B., D.K.), a data consultant (T.M.), and another genetic counselor (M.C.) (Table S1).

<u>Operational definition of actionability.</u> Because the NBSxWGS OTA specified that screened conditions must be actionable within the first year of life, the BEACONS-NBS condition list committee first developed an operational definition of “actionable.” Actionable was defined as a clinically-available intervention or surveillance strategy that could be initiated in infancy and expected to meaningfully alter medical management and improve health outcomes. Qualifying actions included therapeutic diets, medications, and definitive treatments such as hematopoietic stem cell transplantation or gene-based therapies, as well as interventions to prevent harmful exposures (e.g., avoidance of ionizing radiation or live vaccines), and surveillance for condition-related sequelae. As part of the BEACONS-NBS study, participants with positive screening results will receive a list of relevant local specialists, but conditions were considered actionable regardless of whether the recommended interventions are available in every region.

Certain forms of utility were not considered sufficient, in isolation, to meet the definition of actionability. These included avoiding a diagnostic odyssey; access to participation in clinical trials or to experimental therapies; initiation of supportive services (e.g., physical, occupational, or speech therapy); and personal, familial, psychosocial, or economic benefits, such as informing reproductive decision-making or facilitating cascade testing for parents.

<u>Inclusion criteria.</u> Conditions were eligible for inclusion if they met three criteria derived from the 2025 consensus recommendations of the International Consortium on Newborn Sequencing (ICoNS)^22^ (Table 1). Each condition was required to have: (1) an approved treatment or published surveillance guideline that was recommended to begin before one year of age; (2) a non-genetic diagnostic marker (e.g., laboratory result or characteristic imaging finding) or low-risk surveillance strategy; and (3) reliable detection of likely pathogenic and pathogenic (LP/P) variants in the causative gene using short-read WGS.

**Table 1.** Inclusion criteria for genetic conditions in BEACONS-NBS.

|  |  |
| --- | --- |
| 1. | Association with a clinically-approved treatment or published surveillance guideline that could be initiated before one year of age and is expected to substantially reduce condition severity. |
| 2. | Association with supportive diagnostic features beyond genetic testing, such as a laboratory biomarker, imaging finding, physical sign, or clinical diagnostic criterion, that is expected to be present prior to the initiation of treatment. For conditions without a suggestive non-molecular biomarker or examination finding, the surveillance or treatment strategy should be generally considered to be of low risk and potentially high benefit (e.g., abdominal ultrasounds to screen for embryonal tumors in a hereditary cancer predisposition syndrome). |
| 3. | The capacity for short-read whole genome sequencing to reliably detect likely pathogenic or pathogenic variants. |

Regarding the third criterion, GeneDx, the laboratory performing sequencing in BEACONS-NBS, confirmed that at least 90% of each gene would have a minimum read depth of 15x. They excluded genes with known pseudogenes or significant paralog homology that could compromise variant detection. In select cases, genes were included even if the most common LP/P variant type could not be detected by short-read WGS (e.g., the common inversion associated with *F8*-related hemophilia A), given the potential to identify other deleterious variants. Variant classification will be carried out in accordance with criteria set by the guidelines by the American College of Medical Genetics (ACMG) and Association for Molecular Pathology,^23^ with modifications for individual conditions as recommended by the Clinical Genome Resource Variant Curation Expert Panels.^24^

<u>Consideration of prevalence.</u> In accordance with consensus recommendations from ICoNS,^22^ prevalence was not used to determine the inclusion of conditions. Both conditions with high prevalence (e.g.,*G6PD*-related glucose-6-phosphate dehydrogenase deficiency) and ultra-rare conditions were included.

<u>Consideration of penetrance.</u> Penetrance (i.e., the likelihood that a person with an LP/P variant will develop symptoms of the condition) was not explicitly considered in the inclusion criteria.

This decision reflects the current lack of reliable data regarding penetrance for most gene-condition relationships. When possible, we excluded variants and genes for which reported penetrance was so low that the variant-condition or gene-condition relationship was uncertain (e.g., *DUOX2,* HGNC:13273).^25^

<u>Consideration of age of actionability.</u> Conditions with a variable age of onset were included if a severe infantile presentation was known, even when later-onset phenotypes also exist.

Additionally, some conditions with hallmark clinical features that typically manifest after the first year of life were included if interventions or surveillance initiated in infancy could reduce later morbidity. For example, infants at risk for *NF1*-related neurofibromatosis are recommended to have tibial radiographs to identify osseous lesions before weight-bearing, allowing earlier management and prevention of complications.^26^ Variants or modes of inheritance that are associated with attenuated or adult-onset forms of conditions will not be reported (Table S2).

*Development of the BEACONS-NBS list of genetic conditions for screening implementation* <u>Draft 1.</u> The initial list of conditions (Table S3) was assembled by aggregating genes from three sources (Figure 1).

**Figure 1.**
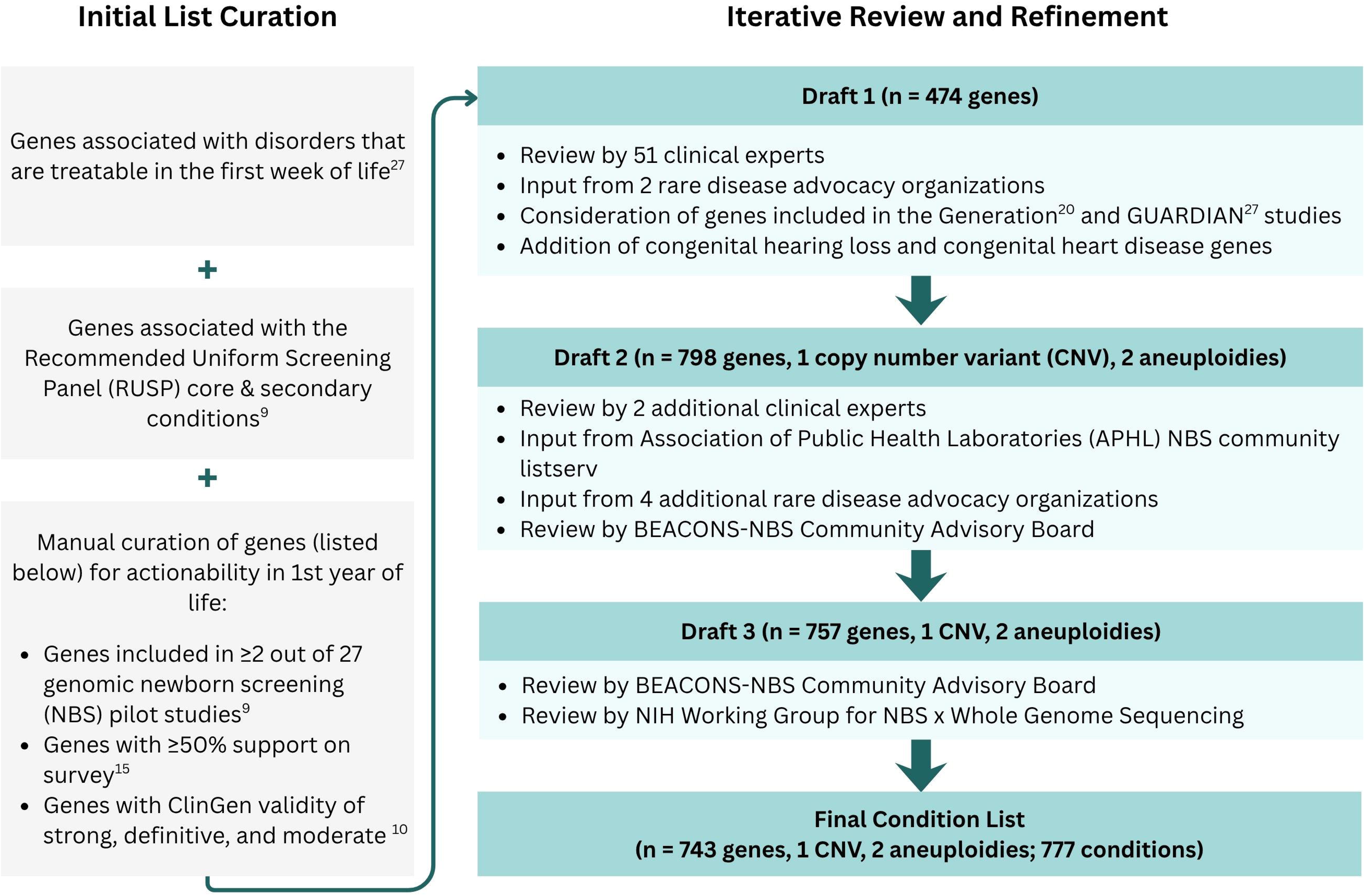
BEACONS-NBS condition list curation process. Key informants suggested the iterative addition and removal of genes as each draft was developed. Of note, genes associated with conditions not actionable in the first year of life, genes with insufficient evidence of gene–disease relationship, and genes not technically feasible for short-read WGS were excluded at each stage.

First, 267 genes associated with conditions that are actionable within the first week of life were added.^27^ Second, we evaluated genes that both appeared in at least two genomic NBS studies or for-profit panels^9^ and received ≥50% support in a survey of rare disease experts^15^ (n = 246).

These genes were then manually reviewed to confirm alignment with BEACONS-NBS inclusion criteria; 143 were included. Third, at the request of prospective state and territorial PHLP partners, we incorporated 103 genes associated with core and secondary conditions currently on the Recommended Uniform Screening Panel (RUSP), compiled from an analysis of genomic NBS programs^9^ and supplemented with internal data from GeneDx. We did not initially include genes related to hearing loss or congenital heart disease, although these were added in later drafts. After combining these three lists, 39 duplicate genes were removed, leading to a total of 474 genes.

<u>Draft 2.</u> To create Draft 2 (Table S4), we incorporated feedback from clinical experts (Table S5), rare disease advocacy organizations, and GeneDx, and reviewed other genomic NBS studies.

We engaged 51 clinical experts who evaluated genes in Draft 1 within their areas of expertise. Twenty-one clinical experts were contacted directly by the BEACONS-NBS condition list committee; they invited 29 colleagues to collaborate. One clinical expert contacted the BEACONS-NBS team directly to review the condition list. The Clinical Immunology Society (CIS) NBS x Genome Sequencing (GS) task force (led by X.P.P., M.J.K., J.D.M., and J.M.P.) created their own committee to revise the BEACONS-NBS list of inborn errors of immunity (IEI).

Clinical experts recommended removal of conditions that did not meet BEACONS-NBS inclusion criteria (Table S6) and proposed additional conditions, which were then evaluated by the BEACONS-NBS condition list committee. Of note, clinical experts reevaluated all secondary RUSP conditions to determine that they met BEACONS-NBS inclusion criteria, and 19 of these genes, including *HBA1* (HGNC:4823) and *DNAJC19* (HGNC:30528), were removed (Table S7). Nineteen genes related to congenital heart disease and 12 genes related to hearing loss were added.

All of the IEI genes in Draft 1 were reevaluated and replaced with a new list of genes proposed by the CIS NBS x GS task force. After removing duplicates with other clinical areas and excluding genes that did not meet BEACONS-NBS inclusion criteria, 236 IEI genes were included in Draft 2.

In total, 341 genes were added and 22 genes were removed between Drafts 1 and 2 due to the recommendations of clinical experts.

Representatives of two rare disease advocacy organizations, the Cure GM1 Foundation and the Plasminogen Deficiency Foundation, contacted the BEACONS-NBS condition list committee directly with suggestions, leading to the addition of one gene.

A review of the genes from the Generation study^11^ and Group 1 of the Genomic Uniform-screening Against Rare Disease in All Newborns (GUARDIAN) genomic NBS study,^28^ which contains treatable conditions, was initiated, yielding the addition of 37 genes in Draft 2.

As Draft 2 was developed, GeneDx initiated a continuous review of the condition list and recommended the removal of 14 genes due to technical limitations of short-read WGS (1 with insufficient coverage, 3 with pseudogenes, 5 with paralog homology, and 5 with other limitations detailed in Table S8) and 17 genes that did not meet clinical criteria due to limited evidence of a gene–disease relationship (Table S6). Draft 2 included a total of 798 genes, one copy number variant (CNV), and two aneuploidies.

<u>Draft 3.</u> To design Draft 3 (Table S9), we incorporated feedback from two additional clinical experts, the Association of Public Health Laboratories (APHL) NBS community listserv, and four national rare disease advocacy organizations (EveryLife Foundation, Genetic Alliance, Global Genes, and National Organization for Rare Disorders). We also continued to evaluate genes from other genomic NBS studies and conducted an ongoing review of the suitability of WGS for detecting LP/P variants in these conditions.

In total, these groups recommended adding 26 genes, 21 of which were incorporated into Draft 3. They also recommended removing 11 genes that did not meet BEACONS-NBS inclusion criteria, all of which were subsequently removed. The remaining genes from the Generation study and Group 1 of the GUARDIAN study also underwent continued review, resulting in the addition of 32 more genes. GeneDx continued their evaluation of the condition list, resulting in the removal of an additional 10 genes due to technical limitations and 72 genes which did not meet clinical inclusion criteria (Table S6).

During the development of Draft 3, Draft 2 was shared with the co-chairs of the BEACONS-NBS Community Advisory Board (CAB) (A.M.G. and N.B.), a committee which is made up of multidisciplinary members within and external to the NBS community, including parents, community advocates, bioethicists, clinical genomics experts, NBS leaders, and others. Draft 2 was also shared with the APHL NBS community listserv, which includes all state and territorial NBS and follow-up programs, corporate members, clinicians, family advocates, and other NBS partners. The BEACONS-NBS condition list committee received detailed feedback from two PHLPs (Puerto Rico and Oregon), who recommended the addition of 10 genes, five of which were incorporated into Draft 3, which in total includes 757 genes, one CNV, and two aneuploidies.

<u>Draft 4.</u> To create Draft 4, the BEACONS-NBS condition list committee incorporated feedback from the NIH Working Group (WG) for NBSxWGS and the CAB.

As required by the NIH OTA, the NIH WG for NBSxWGS conducted a detailed review of Draft 3. They suggested the addition of one gene (*NF1,* HGNC:7765) and the removal of six genes for which they felt that treatment does not substantially change the natural history of the condition (Table S6). The BEACONS-NBS condition list committee also added two additional genes which met inclusion criteria.

At this stage, the BEACONS-NBS condition list committee also transitioned from a gene-based dataset, in which each row represented a gene, to a condition-based dataset, in which each row represented a condition. This change in the organization of the dataset reflects both the practical nuances of WGS analysis, in which many genes give rise to disparate phenotypes with differing modes of inheritance or mechanisms of action, and the interests of prospective participants, who are more likely to consider the conditions for which their child may be screened than the genes associated with these conditions.

The BEACONS-NBS condition list committee also participated in discussions with the BEACONS-NBS CAB about whether conditions primarily managed through surveillance, such as hereditary cancer predisposition syndromes, should be offered as an optional category that parents could elect separately from conditions with established treatment interventions. The CAB recommended keeping the condition list organized as a single group; this recommendation was adopted. Draft 4 contains 743 genes, one CNV, and two aneuploidies.

<u>BEACONS-NBS condition list.</u> The NIH WG for NBSxWGS reviewed and approved Draft 4 for implementation. This list is henceforth referred to as “the BEACONS-NBS condition list,” which will be reevaluated and revised at the halfway point of the study.

### Organization of the BEACONS-NBS condition list

An interactive interface showing the BEACONS-NBS condition list has been published on the BEACONS-NBS website (https://www.beaconsnbs.org/condition-list) (Figure 3). For each condition, 13 attributes which describe the condition and justify its inclusion, are listed: Hugo Gene Nomenclature gene symbol; reporting name of the condition, derived from dyadic naming methods developed by ClinGen^10^ and GeneDx; clinical area; disease area; Online Mendelian Inheritance in Man number^29^; inheritance pattern of variants that will be reported; disease mechanism of the variants that will be reported; a brief condition description; treatments or surveillance strategies collected from RxGenes^6^ and literature review; a rationale for early detection; non-genetic signs, symptoms, and biomarkers supporting diagnosis; citations; and specific reporting information for select conditions (see Supplementary Methods for further details).

### Descriptive statistics

Characteristics of the BEACONS-NBS condition list were tabulated and expressed as descriptive statistics.

### Comparison with other gene and condition lists

We determined the intersection between the genes associated with the BEACONS-NBS condition list and the genes in other major genomic NBS research studies, including the Generation Study,^11^ Early Check,^12^ and Group 1 of the GUARDIAN study^28^. We also determined the intersection between the genes on the BEACONS-NBS condition list and those included in commercial carrier screening panels, including the Myriad Foresight Carrier Screen^30^ and Natera Horizon 421, and the larger Natera Horizon 835 and LabCorp 790 PLUS Carrier panels.^31^

## Results

### Descriptive statistics

The BEACONS-NBS condition list includes 743 genes, one CNV, and two aneuploidies associated with 777 genetic conditions (Figure 2, Table S10). In total, 555 conditions (71.4%) are autosomal recessive, 169 (21.8%) are autosomal dominant, 49 (6.3%) are X-linked, and 1 (0.1%) is Y-linked (Figure S1). One condition (0.1%), DiGeorge Syndrome, is due to a CNV and two conditions (0.3%), Down syndrome and Turner syndrome, are due to aneuploidies.

**Figure 2.**
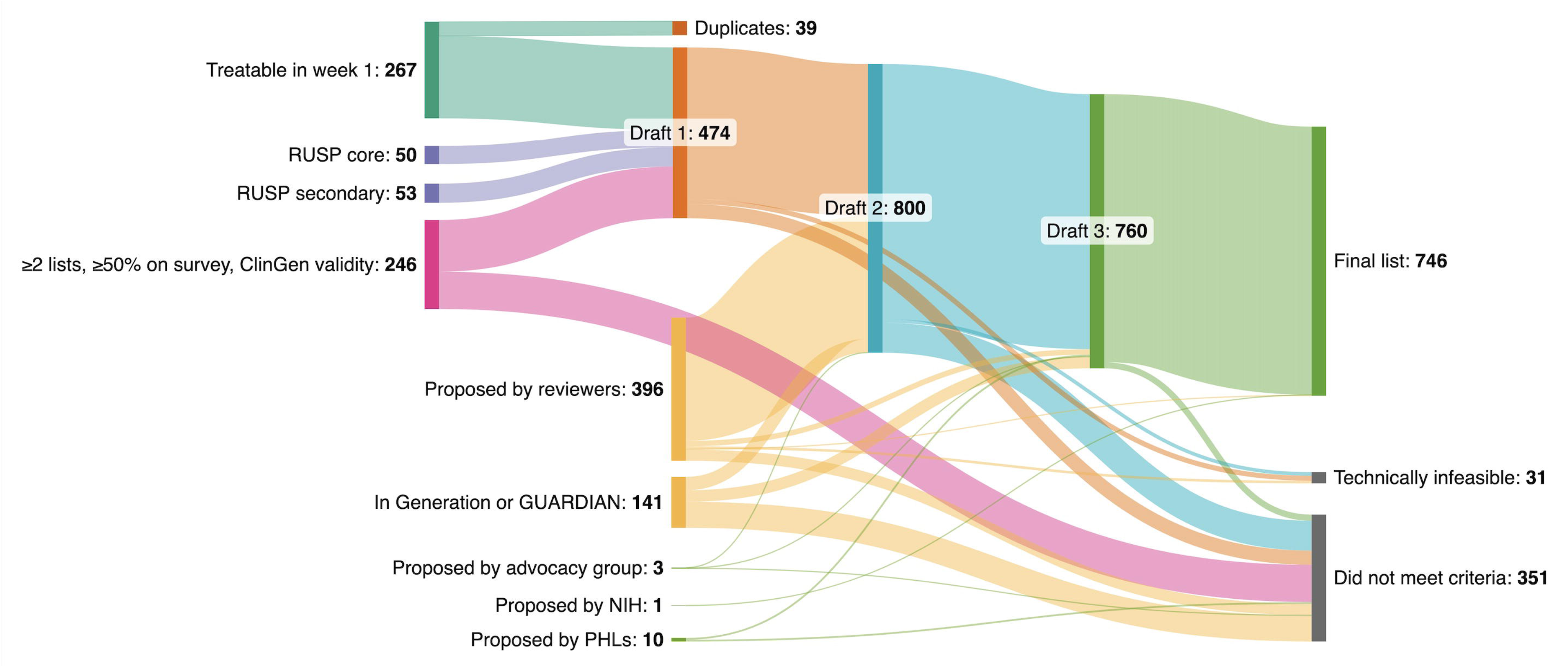
Aggregation and review of genes associated with conditions included in the BEACONS-NBS condition list. Vectors show the development of the BEACONS-NBS condition list (Drafts 1-4).

**Figure 3.**
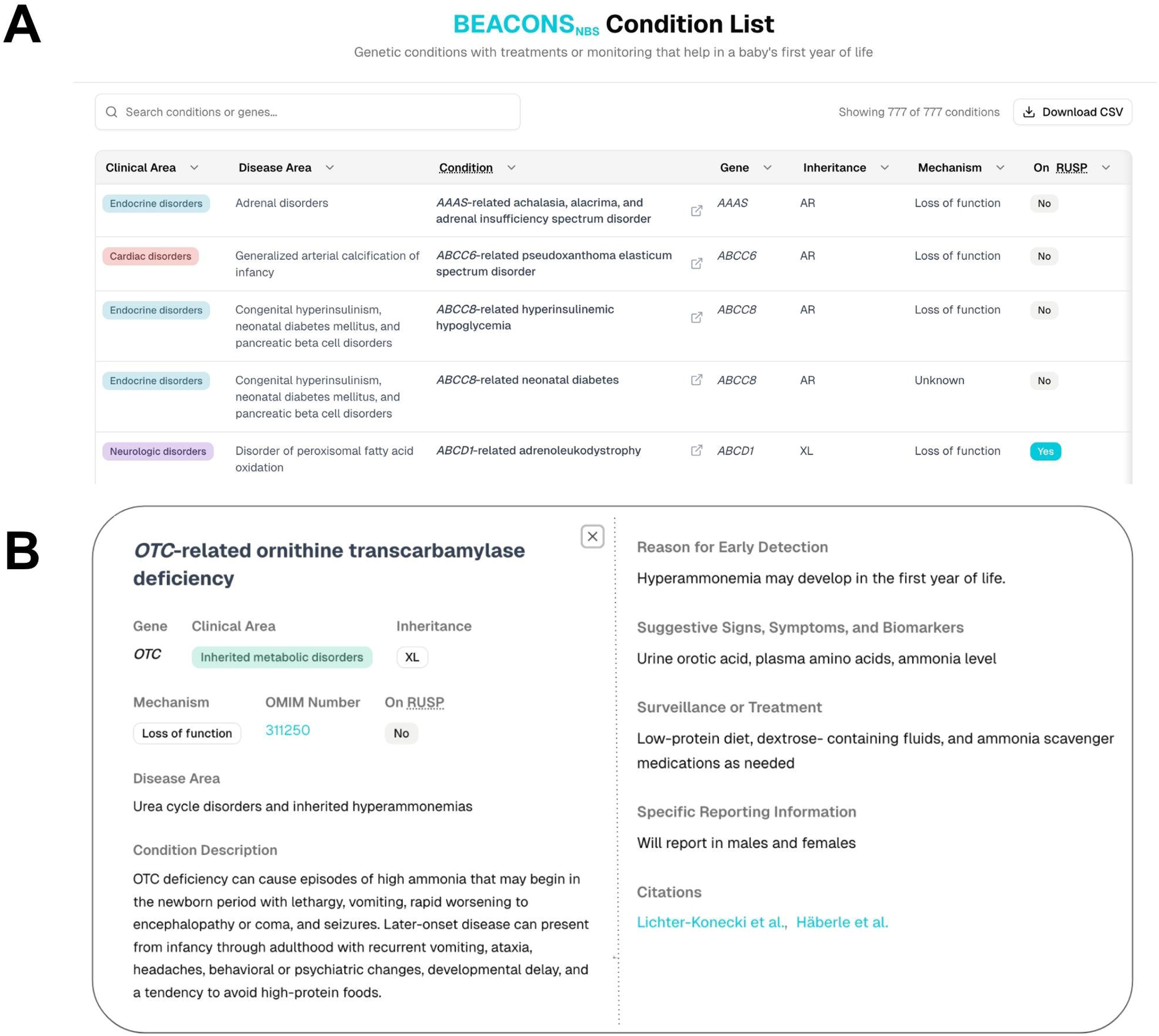
BEACONS-NBS condition list interactive interface. (A) Main table view displaying each condition with its associated gene, clinical area (the medical specialty most likely to manage the primary treatment for condition), inheritance pattern, mechanism, and RUSP status. Users can search, filter, and download the full list as a CSV file. (B) Expanded condition view for an individual entry (*OTC*-related ornithine transcarbamylase deficiency) showing additional information, including condition description; rationale for early detection; suggestive signs, symptoms, and biomarkers; surveillance or treatment; and citations that describe early treatment of the condition.

Together, IEI (289 conditions, 37.2%), inherited metabolic disorders (IMD) (146 conditions, 18.7%), and endocrine conditions (141 conditions, 18.1%) account for over 70% of the list (Figure S1). Only 106 of 777 conditions (13.6%) are currently core conditions on the RUSP.

Nearly all conditions on the BEACONS-NBS condition list (725 conditions, 93.3%) have at least one associated non-genetic marker that could support diagnosis following a positive genomic screen. The 52 conditions without a non-genetic marker (Table S11), 19 of which are hereditary cancer predisposition syndromes, either have published clinical diagnostic criteria or were included because the BEACONS-NBS conditions list committee considered the recommended treatment or surveillance to be of sufficiently low risk and high benefit.

### Comparison with other genomic NBS research studies

The BEACONS-NBS condition list includes a greater number of genes than those in the Generation study, Early Check, or Group 1 of the GUARDIAN genomic NBS research study (Figure 4). The BEACONS-NBS condition list shares 369 genes with the Generation study. Among BEACONS-NBS and the two other U.S.-based programs (Early Check and Group 1 of the GUARDIAN study), 126 genes were shared.

**Figure 4.**
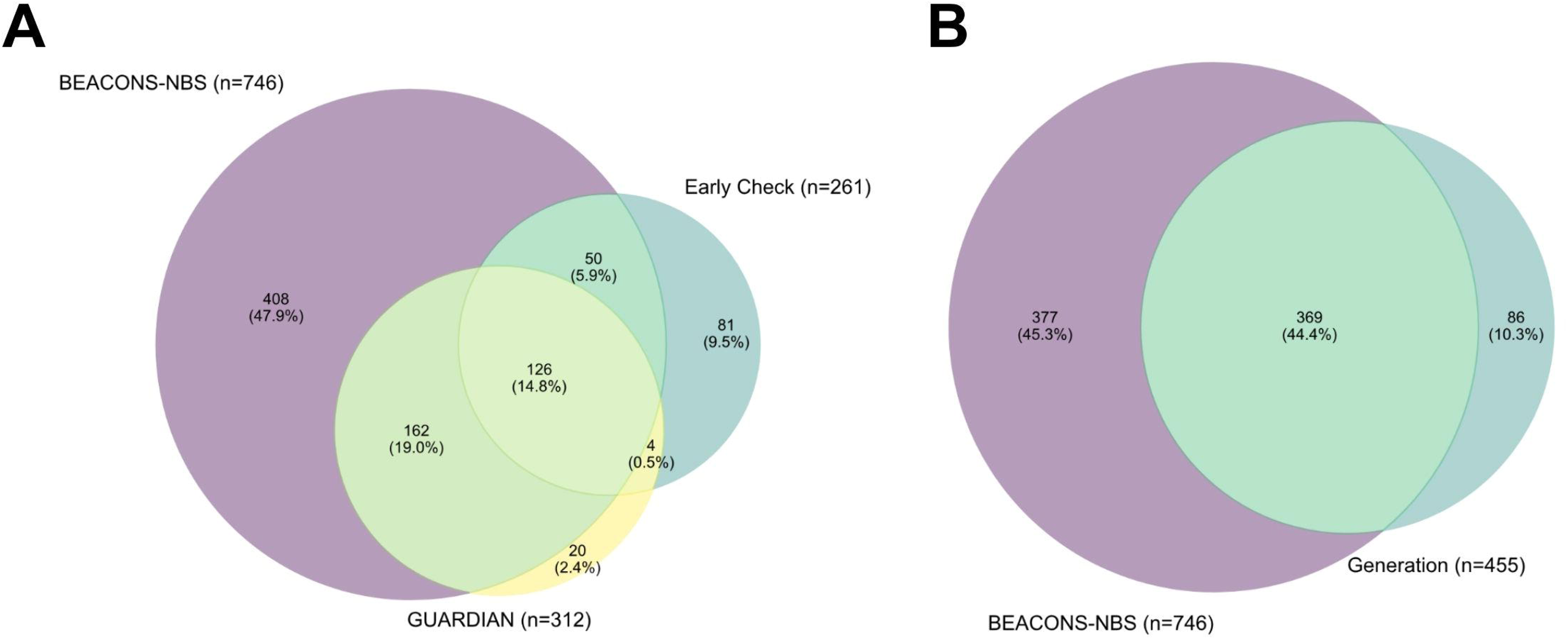
Intersection of the BEACONS-NBS condition list with other genomic newborn screening research programs. (A) Comparison of genes associated with the BEACONS-NBS condition list with two U.S.-based programs (Early Check and Group 1 of the GUARDIAN study) showing shared and unique genes across lists. (B) Comparison of genes associated with the BEACONS-NBS condition list with the Genomics England Generation study gene list. Gene list from BabySeq (4,299 genes) is not shown.

BEACONS-NBS includes 280 genes that were in neither Early Check, Group 1 of the GUARDIAN study nor in the Generation study. Most were related to IEI (n=128, 45.7%), endocrine disorders (n=52, 18.6%) and IMD (n=35, 12.5%) (Table S12).

### Comparison with expanded carrier screening panels

BEACONS-NBS shares a total of 180 genes with the Myriad Foresight Carrier Screen and/or Natera Horizon 421 expanded carrier screening panel. BEACONS-NBS shares 313 genes with the Natera Horizon 835 and/or LabCorp 790 PLUS Carrier panels. However, the majority of genes associated with the BEACONS-NBS condition list (n = 433/746, 58%) were not on any of these carrier screening panels (Figure S2).

## Discussion

### Alignment of the BEACONS-NBS condition list with the Wilson-Jungner principles

BEACONS-NBS will evaluate the feasibility of incorporating genomic sequencing for 777 conditions into PHLPs’ NBS workflows. Although the BEACONS-NBS study will greatly expand the breadth of NBS in participating programs, the condition list was designed to remain broadly consistent with key Wilson–Jungner principles for population screening.^3^ Specifically, included conditions are associated with important health outcomes, have an identifiable early stage that can be confirmed through clinical evaluation or diagnostic testing, and have established treatments or surveillance strategies that can be initiated in infancy.

At the same time, implementation of the BEACONS-NBS condition list will also challenge some of the Wilson-Jungner principles. For example, these principles suggest that there should be an agreed-upon policy on who to treat as a patient and local facilities available for diagnosis and treatment. Although BEACONS-NBS will provide ACTion sheets in the style of those produced by the ACMG^32^, standardized approaches to the management of infants with positive genomic screening results have not yet been established. In addition, access to diagnostic services and specialized treatment resources may vary across the seven participating states and territories. Evaluation of long-term clinical outcomes and the economic implications of genomic screening are beyond the primary scope of BEACONS-NBS but are important priorities for future research.

### Comparison of BEACONS-NBS to other leading genomics initiatives

Although the BEACONS-NBS list is larger than those used by other U.S. and U.K. genomic NBS programs, many conditions are shared across them, reflecting a common emphasis on early-onset, medically actionable conditions. Each program also includes a distinct subset of genes, suggesting differences in inclusion criteria, particularly age of actionability, and operational interpretations of actionability. In particular, at the direction of the CIS NBS x GS task force, BEACONS-NBS includes more genes associated with IEIs than other studies. This in part reflects the program’s broad operational definition of actionability, which encompasses infection-prevention strategies and modification of routine childhood care, such as avoidance of live vaccines, rather than just targeted medical treatments.

The BEACONS-NBS condition list also differs from large carrier screening panels, which are designed to inform reproductive decision-making.^33^ Carrier screening panels often include conditions without clinically-available treatments, as well as conditions selected due to higher prevalence in specific populations; these selection principles do not align with BEACONS-NBS inclusion criteria.^34,35^ In contrast, BEACONS-NBS prioritizes potential clinical benefit to infants in the first year of life, and includes 169 autosomal dominant conditions that are usually excluded from carrier screening. Consequently, parents who have already undergone carrier screening may still receive new, clinically-relevant information about their infant through BEACONS-NBS.

### Key challenges and limitations in condition list development

One challenge of developing the BEACONS-NBS condition list was operationalizing “actionability,” a term which has disparate qualitative and quantitative definitions.^13,36–39^ Although some parents have expressed interest in genomic NBS for conditions for which only supportive care is currently available,^18,28^ we prioritized clinical utility, with an emphasis on therapeutic efficacy.^38^ This approach will not shorten the diagnostic odyssey for children with conditions that have only supportive treatments, nor will it enable parents of presymptomatic children to pursue experimental treatments when no clinical treatment is available. In BEACONS-NBS, parents and clinicians can request reanalysis of WGS in infants who later develop signs of a genetic condition, which will help mitigate the extent of this limitation. The inclusion criteria for conditions will be revisited at the halfway point of the study and there will be an opportunity to potentially broaden or narrow the conditions that are included.

Another challenge was addressing penetrance and variability in age of onset. We did not select conditions based on estimated penetrance because reliable data on this topic are lacking for most conditions, particularly when individuals with LP/P variants are identified via population screening.^40^ Many conditions lack well-defined genotype-phenotype correlations, leading to a wide range of severity and age of onset. To maximize sensitivity, we included conditions for which a severe infantile presentation has been described, even when onset may occur later in childhood or adolescence. For some genes, such as *RET*, reporting will be limited to specific variants associated with infantile-onset symptoms. Consistent with conventional NBS, we also included conditions that may present in the first days of life, before results may be returned. This approach recognizes that age of onset is often variable and genomic information may still guide treatment decisions in the first weeks or months of life, particularly in attenuated cases. Over time, prospective studies that link genotypic and detailed phenotypic information, including BEACONS-NBS, will help address knowledge gaps related to genomic penetrance, variable expressivity, and age of symptom onset, thereby informing the clinical utility of this screening modality.

Assembling a list of conditions which are actionable in the first year of life is an inductive process with several limitations. Because no definitive database describes the technical and clinical attributes relevant to genomic NBS for every gene, we developed a new dataset tailored to our objectives. Given that conditions were added iteratively by multiple key informants, some conditions that meet the BEACONS-NBS inclusion criteria may have been unintentionally omitted. Another important issue, highlighted by the BEACONS-NBS CAB, is whether screening should be limited to LP/P variants or expanded to include pathogenic-leaning variants of uncertain significance (VUS) in order to improve detection among individuals of non-European ancestry. This consideration must be balanced against the potential risk that reporting such variants could increase the identification of low penetrance or ultimately benign variants in underrepresented populations.^41^ The BEACONS-NBS team decided that, at the outset of the study, VUS will not be included,but approximately one year after participant recruitment begins, there will be an opportunity to revise reporting strategies. The lack of long-read WGS is also a limitation that resulted in the removal of several genes that have associated pseudogenes or paralogs, such as *CYP21A2* (HGNC:2600), associated with classic congenital adrenal hyperplasia.

## Conclusion

The BEACONS-NBS condition list represents an effort to apply the historic Wilson-Jungner principles to genomic NBS. It reflects both areas of consensus with other genomic newborn screening research programs and important differences arising from cultural perspectives, variation in inclusion criteria related to age of symptom onset, differing operational definitions of actionability, uncertainty regarding variable expressivity and penetrance, and evolving genotype–phenotype knowledge. Rather than a definitive catalog, the list should be viewed as a dynamic framework that will be refined as new empirical data, technical improvements, stakeholder input, and targeted therapies emerge. By prospectively evaluating the implementation of genomic NBS across several PHLPs, BEACONS-NBS will generate much-needed evidence about the feasibility, clinical utility, acceptability, and limitations of this screening modality. These findings may inform policy discussions about how genomic information can be incorporated ethically and effectively into public health NBS programs as diagnostic capabilities, therapies, and societal expectations continue to evolve.

## Data Availability

All data produced in the present work are contained in the manuscript

## Acknowledgments

BEACONS-NBS is carried out as a collaborative study supported by the National Institutes of Health, OTA 1OT2OD040029-01. The authors thank the staff and participants of BEACONS-NBS for their important contributions. We also thank GeneDx and Illumina for their collaboration and support. RCG thanks the Franca Sozzani Fund and Coleman Family for support. We also thank Dr. PJ Brooks, Dr. Melissa Parisi, Dr. George Papanicolaou, Dr. Mollie Minear, Dr.

Kathleen Huntzicker, and Ms. Linda Ho, members of the NIH Working Group for NBSxWGS, for insightful discussions and helpful feedback, and Dr. Elliot Kozin for contributing his expertise on hearing loss disorders for inclusion on the condition list.

## Data availability

The published article includes all datasets generated or analyzed during this study.

## Author Contributions

*Conceptualization:* N.B.G., I.A.H., M.P.W., R.C.G.

*Data curation:* N.B.G., B.A.J., H.S., A.B., E.B., K.G.L., R.Z., H.M.M., T.E., A.K., I.A.H., R.S.A.,

I.A., M.B., C.J.B., M.J.B., S.W.C., M.C., Y.-M.C., S.C., S.C.C.C., O.M.D., L.R.D., L.D., J.F., A.F.,

R.D.G., J.G., R.G.-M., E.G., R.C.H., A.H., F.H., A.M.H., C.J., M.J.K., M.K., V.K., R.J.K., B.S.L.,

A.E.L., J.A.M., R.A.M., D.T.M., J.D.M., D.M.M., B.M., L.D.N., X.P.P., E.M.P., C.D.P., S.E.P.,

S.R.P., C.M.P., J.M.P., A.E.R., V.G.S., D.M.S., C.M.S., A.S., K.E.S., W.T., T.K.T., J.R.T., M.A.W., A.J.W., H.Z.B.

*Formal analysis:* T.M., H.S.

*Funding acquisition:* R.C.G., N.B.G., I.A.H., M.P.W., C.B.Z., S.A.C., B.A.J., S.S. (Sikha Singh), J.O., S.P., A.J.G.

*Investigation:* N.B.G., B.A.J.

*Methodology:* N.B.G., B.A.J.

*Project administration:* N.B.G., B.A.J., S.A.C., C.B.Z., H.S., A.G., N.B., J.O., S.S. (Sikha Singh), G.Z.

*Resources:* B.A.J., A.B., E.B., K.G.L., R.Z., H.M.M., T.E., A.K.

*Software:* S.S. (Shreyas Srinath), H.S., N.B.G.

*Supervision:* N.B.G., B.A.J., A.M.G., N.B., J.O., S.S. (Sikha Singh), G.Z., S.P., A.J.G., I.A.H., M.P.W., R.C.G.

*Visualization:* T.M., N.B.G., H.S.

*Validation:* T.M., A.V.M.

*Writing-original draft:* N.B.G., H.S.

*Writing-review & editing:* All authors

## Ethics Declaration

As no individualized or patient-specific data were used, Institutional Review Board (IRB) or Research Ethics Committee (REC) approval were not required.

## Declaration of generative AI and AI-assisted technologies in the manuscript preparation process

During the preparation of this work the authors used ChatGPT in order to improve upon paragraph organization, sentence syntax, and grammar, and identify references on various relevant topics. After using this tool/service, the authors reviewed and edited the content as needed and take full responsibility for the content of the published article.

## References

1. Watson MS, Lloyd-Puryear MA, Howell RR. The progress and future of US newborn screening. Int J Neonatal Screen. 2022;8(3):41.

2. Gaviglio A, McKasson S, Singh S, Ojodu J. Infants with congenital diseases identified through newborn screening-United States, 2018-2020. Int J Neonatal Screen. 2023;9(2):23.

3. Wilson JMG, Jungner G. Principles and Practice of Screening for Diseases. World Health Organization; 1968.

4. Gramer G, Hoffmann GF. Second-tier strategies in newborn screening - potential and limitations. Med Genet. 2022;34(1):21–28.

5. Kucera KS, Taylor JL, Robles VR, et al. A voluntary statewide newborn screening pilot for spinal muscular atrophy: Results from Early Check. Int J Neonatal Screen. 2021;7(1):20.

6. Bick D, Bick SL, Dimmock DP, Fowler TA, Caulfield MJ, Scott RH. An online compendium of treatable genetic disorders. Am J Med Genet C Semin Med Genet. 2021;187(1):48–54.

7. Schnabel-Besson E, Mütze U, Dikow N, et al. Wilson and Jungner revisited: Are screening criteria fit for the 21st century? Int J Neonatal Screen. 2024;10(3):62.

8. Stark Z, Scott RH. Genomic newborn screening for rare diseases. Nat Rev Genet. 2023;24(11):755–766.

9. Minten T, Bick S, Adelson S, et al. Data-driven consideration of genetic disorders for global genomic newborn screening programs. Genet Med. 2025;27(7):101443.

10. Rehm HL, Berg JS, Brooks LD, et al. ClinGen--the clinical genome resource. N Engl J Med. 2015;372(23):2235–2242.

11. Kaplanis J, Deen D, Sivakumar P, et al. Assessment of the variant prioritization strategy for genomic newborn screening in the Generation Study. Genet Med. 2025;27(10):101532.

12. Cope HL, Milko LV, Jalazo ER, et al. A systematic framework for selecting gene-condition pairs for inclusion in newborn sequencing panels: Early Check implementation. Genet Med. 2024;26(12):101290.

13. Hunter JE, Jenkins CL, Bulkley JE, et al. ClinGen’s Pediatric Actionability Working Group: Clinical actionability of secondary findings from genome-scale sequencing in children and adolescents. Genet Med. 2022;24(6):1328–1335.

14. Downie L, Halliday J, Lewis S, Amor DJ. Principles of genomic newborn screening programs: A systematic review. JAMA Netw Open. 2021;4(7):e2114336.

15. Gold NB, Adelson SM, Shah N, et al. Perspectives of rare disease experts on newborn genome sequencing. JAMA Netw Open. 2023;6(5):e2312231.

16. Etchegary H, Dicks E, Hodgkinson K, Pullman D, Green J, Parfey P. Public attitudes about genetic testing in the newborn period. J Obstet Gynecol Neonatal Nurs. 2012;41(2):191–200.

17. Timmins GT, Wynn J, Saami AM, Espinal A, Chung WK. Diverse parental perspectives of the social and educational needs for expanding newborn screening through genomic sequencing. Public Health Genomics. 2022;25(5-6):1–8.

18. Gold NB, Omorodion JO, Del Rosario MC, et al. Preferences of parents from diverse backgrounds on genomic screening of apparently healthy newborns. J Genet Couns. 2025;34(2):e1994.

19. Joseph G, Chen F, Harris-Wai J, Puck JM, Young C, Koenig BA. Parental views on expanded newborn screening using whole-genome sequencing. Pediatrics. 2016;137 Suppl 1(Supplement):S36–S46.

20. Downie L, Bouffler SE, Amor DJ, et al. Gene selection for genomic newborn screening: Moving toward consensus? Genet Med. 2024;26(5):101077.

21. Betzler IR, Hempel M, Mütze U, et al. Comparative analysis of gene and disease selection in genomic newborn screening studies. J Inherit Metab Dis. 2024;47(5):945–970.

22. Downie L, Yeo J, Minten T, et al. Operationalizing the Wilson-Jungner principles for the genomics era: Consensus recommendations from the International Consortium on Newborn Sequencing. Genet Med. 2026;28(1):101618.

23. Richards S, Aziz N, Bale S, et al. Standards and guidelines for the interpretation of sequence variants: a joint consensus recommendation of the American College of Medical Genetics and Genomics and the Association for Molecular Pathology. Genet Med. 2015;17(5):405–423.

24. Rivera-Muñoz EA, Milko LV, Harrison SM, et al. ClinGen Variant Curation Expert Panel experiences and standardized processes for disease and gene-level specification of the ACMG/AMP guidelines for sequence variant interpretation. Hum Mutat. 2018;39(11):1614–1622.

25. Peters C, Nicholas AK, Schoenmakers E, et al. DUOX2/DUOXA2 mutations frequently cause congenital hypothyroidism that evades detection on newborn screening in the United Kingdom. Thyroid. 2019;29(6):790–801.

26. Miller DT, Freedenberg D, Schorry E, et al. Health supervision for children with neurofibromatosis type 1. Pediatrics. 2019;143(5):e20190660.

27. Cohen JL, Duyzend M, Adelson SM, et al. Advancing precision care in pregnancy through a treatable fetal findings list. Am J Hum Genet. 2025;112(6):1251–1269.

28. Ziegler A, Koval-Burt C, Kay DM, et al. Expanded newborn screening using genome sequencing for early actionable conditions. JAMA. 2025;333(3):232–240.

29. Amberger JS, Bocchini CA, Schiettecatte F, Scott AF, Hamosh A. OMIM.org: Online Mendelian Inheritance in Man (OMIM®), an online catalog of human genes and genetic disorders. Nucleic Acids Res. 2015;43(Database issue):D789–D798.

30. Plan and prepare with prenatal genetic screening from Myriad. Myriad Genetics. July 22, 2024. Accessed February 18, 2026. https://myriad.com/prenatal-genetic-screening/

31. Comprehensive Screening Options from Horizon. Natera. January 3, 2021. Accessed February 18, 2026. https://www.natera.com/womens-health/horizon-advanced-carrier-screening/what-it-screens/

32. *ACMG ACT Sheets and Algorithms*. American College of Medical Genetics and Genomics; 2001.

33. Downie L, Lunke S, Stark Z. The intersection between genetic reproductive carrier screening and genomic newborn screening: Implications for clinical practice. Prenat Diagn. 2025;45(10):1277–1280.

34. Gregg AR, Aarabi M, Klugman S, et al. Screening for autosomal recessive and X-linked conditions during pregnancy and preconception: a practice resource of the American College of Medical Genetics and Genomics (ACMG). Genet Med. 2021;23(10):1793–1806.

35. Grody WW, Thompson BH, Gregg AR, et al. ACMG position statement on prenatal/preconception expanded carrier screening. Genet Med. 2013;15(6):482–483.

36. Lee K, Abul-Husn NS, Amendola LM, et al. ACMG SF v3.3 list for reporting of secondary findings in clinical exome and genome sequencing: A policy statement of the American College of Medical Genetics and Genomics (ACMG). Genet Med. 2025;27(8):101454.

37. Hunter JE, Irving SA, Biesecker LG, et al. A standardized, evidence-based protocol to assess clinical actionability of genetic disorders associated with genomic variation. Genet Med. 2016;18(12):1258–1268.

38. Hayeems RZ, Dimmock D, Bick D, et al. Clinical utility of genomic sequencing: a measurement toolkit. NPJ Genom Med. 2020;5(1):56.

39. Berg JS, Foreman AKM, O’Daniel JM, et al. A semiquantitative metric for evaluating clinical actionability of incidental or secondary findings from genome-scale sequencing. Genet Med. 2016;18(5):467–475.

40. Gelb BD. 2024 ASHG presidential address: Incomplete penetrance and variable expressivity: Old concepts, new urgency. Am J Hum Genet. 2025;112(3):461–466.

41. Chen E, Facio FM, Aradhya KW, et al. Rates and classification of variants of uncertain significance in hereditary disease genetic testing. JAMA Netw Open. 2023;6(10):e2339571.

