## Supplementary appendix for "Selection of Genetic Conditions for Multi-State Genomic Newborn Screening in BEACONS-NBS"

Supplementary Material

TABLE OF CONTENTS

| ***Authorship*** |  |
| --- | --- |
| Supplement A: The Building Evidence and Collaboration for GenOmics in Nationwide Newborn Screening (BEACONS-NBS) team authors | 3 |
| ***Methods*** |  |
| Supplementary Methods | 4 |
| ***Supplemental Tables*** |  |
| Table S1. BEACONS-NBS condition list committee members and their roles | 8 |
| Table S2. Examples of conditions excluded due to onset later than 1 year of life | 9 |
| Table S3. Genes associated with Draft 1 of the BEACONS-NBS condition list (n = 474)* |  |
| Table S4. Genes associated with Draft 2 of the BEACONS-NBS condition list (n = 800)* |  |
| Table S5. List of experts and their clinical specialties | 10 |
| Table S6. Genes proposed but rejected for the BEACONS-NBS condition list for not meeting clinical inclusion criteria (inclusion criteria 1 and 2) | 13 |
| Table S7. Genes associated with secondary RUSP conditions which were excluded from Draft 2 of the BEACONS-NBS condition list | 25 |
| Table S8. Genes proposed but rejected from the BEACONS-NBS condition list due to concerns that detecting likely pathogenic and pathogenic variants would be technically challenging using whole genome sequencing (inclusion criterion 3) | 26 |
| Table S9. Genes associated with Draft 3 of the BEACONS-NBS condition list (n = 760)* |  |
| Table S10. BEACONS-NBS condition list for implementation (n=743 genes, 1 copy number variant, 2 aneuploidies; 777 conditions)* |  |
| Table S11. Conditions on the BEACONS-NBS condition list without a non-genetic suggestive sign, symptom or biomarker | 29 |
| Table S12. Genes associated with the BEACONS-NBS condition list not included in Early Check, Generation, or Group 1 of the GUARDIAN study (n=280)* |  |
| ***Supplemental Figures*** |  |
| Figure S1. Distribution of BEACONS-NBS conditions by clinical area and mode of inheritance | 32 |
| Figure S2. Overlap of genes included in the BEACONS-NBS condition list and commercial carrier screening panels | 33 |

*****Table available in separate excel file

### Supplement A: The Building Evidence and Collaboration for GenOmics in Nationwide Newborn Screening (BEACONS-NBS) team authors

| Swaroop Aradhya | Elizabeth Bair | Amber Begtrup |
| --- | --- | --- |
| David Bick | Carrie L. Blout Zawatsky | Natasha Bonhomme |
| Steven E. Brenner | Elizabeth Butler | Amy Calhoun |
| Danielle Carnival | Megan Clarke | Kenneth Coursey |
| Stephanie A. Coury | Erin Drake | Tessa Ellefson |
| Mario Estevez | Albert Freedman | Amy Gaviglio |
| Nina B. Gold | Aaron Goldenberg | Robert C. Green |
| Nathan Grant | Sue Gullo | Patrice Held |
| Ingrid A. Holm | Justin Hopkin | Britt A. Johnson |
| Dalia Kasperaviciute | Denise Kay | Nicole Kelly |
| Adriann Kern | Brian Kirmse | Katie Kneupper |
| Vilmante Kodyte | Grace Kubin | Katherine G. Langley |
| Zachery Leeker | Anna Lewis | Jennifer Lewis |
| Stuart Lipsitz | D'Andra Luna | Heather M. McLaughlin |
| Dianalee McKnight | Meadhbh Molloy | Brittany L. Nelson |
| Cara O'Neill | Jelili Ojodu | Joe Orsini |
| Katrina Paleologos | Frida Garcia Pelayo | Stacey Pereira |
| Kimberly Piper | Roselle Ponsaran | Deborah Requesens |
| Brendan Reilly | Ledith Resto | Sulay Rivera |
| Virginia Sack | Samantha Sanjeev | Sikha Singh |
| Harini Somanchi | Leslie Soto | Shreyas Srinath |
| Laura Subramanian | Susan Tanksley | Beth Tarini |
| Norma Tavakoli | Page Trotter | Pawan Vohra |
| Melissa P. Wasserstein | Jenifer Waldrop | Kaitlin Weisshappel |
| Sam Woodbury | Guisou Zarbalian | Rebekah Zimmerman |

### Supplementary Methods

*Description of the genetic conditions included in BEACONS-NBS*

Reporting name. Each genetic condition was assigned a “reporting name” corresponding to the specific condition meeting BEACONS-NBS inclusion criteria. Reporting names were derived from a dyadic nomenclature developed by GeneDx, and based on a framework from ClinGen.^1^ The reporting name aligns with the name of the condition which will appear on positive BEACONS-NBS genomic newborn screening reports and corresponding ACTion sheets.

Clinical Area and Disease Area. All conditions were assigned to one of thirteen main clinical areas (cardiac, dermatologic, endocrine, gastrointestinal, hearing loss, hematologic disorders, hereditary cancer predisposition syndromes, inborn errors of immunity, inherited metabolic disorders, neurologic, ophthalmologic, pulmonary, renal) by the BEACONS-NBS condition list committee based on the pediatric specialty who is most likely to manage the primary treatment of each condition. Each condition was also assigned to a disease area subgroup. For inborn errors of immunity, the The Clinical Immunology Society (CIS) NBS x Genome Sequencing (GS) task force used disease groupings based on the The International Union of Immunological Societies (IUIS) classification.^2^ For inherited metabolic disorders, subgroup groupings were extracted from the IEMbase nosology.^3^ Other disease area subgroup designations were assigned by the BEACONS-NBS condition committee and associated clinical experts.

OMIM number. OMIM numbers were obtained directly from the OMIM database. Data was retrieved from the “genemap2.txt” file available at <https://www.omim.org/downloads/>.

Inheritance pattern**.** Inheritance patterns were initially derived from OMIM and subsequently reviewed by clinical experts and the BEACONS-NBS condition list committee. Additional review by GeneDx led to refinements to ensure that reported inheritance aligned specifically with the conditions selected for BEACONS-NBS reporting. Because WGS will be performed on the infant only, conditions with autosomal recessive inheritance will be reported when variants are biallelic. Additional parental studies will be provided by GeneDx to parents of participants to phase variants during infants’ clinical evaluations.

Mechanism. Mechanism of genetic disease information was initially derived from the dyadic nomenclature system developed by GeneDx. These designations were then translated into more widely recognized and accessible mechanism terms for use on the public-facing BEACONS-NBS website. Specifically, GeneDx’s classifications of “neomorph,” “hypermorph,” “antimorph,” and “amorph” were mapped to the corresponding commonly used terms of new function, increased function, dominant negative, and loss of function, respectively.

Condition description. Condition descriptions were initially obtained from GeneDx’s reporting descriptions. These descriptions were then simplified by the BEACONS-NBS condition list committee to improve clarity and readability for a broad audience.

Published treatments, surveillance strategies and rationale for early detection. Information on published treatments and surveillance approaches, as well as the rationale for detection within the first year of life, was compiled through literature review and input from clinical expert reviewers. This information was used to confirm that each condition met BEACONS-NBS actionability criteria.

Suggestive signs, symptoms, and biomarkers. Data on suggestive clinical features, laboratory biomarkers, and other diagnostic indicators were manually curated using action sheets provided by Genomics England, supplemental materials from a study surveying rare disease experts,^4^ and drafted by the BEACONS-NBS condition list committee and clinical experts.

Citations. Citations describing the surveillance and/or treatment for each condition were compiled by the BEACONS-NBS condition list committee and reviewed by clinical experts.

Specific Reporting Information. Reporting information for specific variants is included for a subset of genes with well-established genotype-phenotype correlations. Additionally, for each X-linked condition, it is indicated whether results will be reported for chromosomal males alone or both chromosomal males and females females. Likely pathogenic and pathogenic (LP/P) variants in X-linked conditions will be reported in both chromosomal sexes when an actionable phenotype in females has been previously described in the literature (e.g., *F8*-related hemophilia). Chromosomal sex will be determined as part of the WGS bioinformatic workflow at GeneDx.

**References**

1.	Rehm HL, Berg JS, Brooks LD, et al. ClinGen--the clinical genome resource. *N Engl J Med*. 2015;372(23):2235-2242.

2.	Bousfiha A, Moundir A, Tangye SG, et al. The 2022 update of IUIS phenotypical classification for human Inborn Errors of immunity. *J Clin Immunol*. 2022;42(7):1508-1520.

3.	Lee JJY, Wasserman WW, Hoffmann GF, van Karnebeek CDM, Blau N. Knowledge base and mini-expert platform for the diagnosis of inborn errors of metabolism. *Genet Med*. 2018;20(1):151-158.

4.	Gold NB, Adelson SM, Shah N, et al. Perspectives of rare disease experts on newborn genome sequencing. *JAMA Netw Open*. 2023;6(5):e2312231.

### Table S1. BEACONS-NBS condition list committee members and their roles

| **Name** | **Role** |
| --- | --- |
| Nina B. Gold | Co-chair |
| Britt A. Johnson | Co-chair |
| Ingrid A. Holm | mPI |
| Stephanie A. Coury | Program manager |
| Harini Somanchi | Program manager |
| Katherine G. Langley | GeneDx genetic counselor |
| Amber Begtrup | GeneDx molecular geneticist |
| Rebekah Zimmerman | GeneDx molecular geneticist |
| Thomas Minten | Data consultant |
| Heidi L. Rehm | Consultant |
| David Bick | Consultant |
| Steven E. Brenner | Consultant |
| Dalia Kasperaviciute | Consultant |
| Makena Caron | Genetic counselor research trainee |

### Table S2. Examples of conditions excluded due to onset later than 1 year of life

| **Gene** | **Condition** | **Reason for exclusion** |
| --- | --- | --- |
| *APC* | *APC*-related familial adenomatous polyposis | Although *APC*-related familial adenomatous polyposis is included, the c.3920T>A Ashkenazi Jewish founder variant was excluded due to the recommendation for colon cancer screening to begin at 40 years of age.^1^ |
| *F2* | *F2*-related prothrombin deficiency | While F2-related prothrombin deficiency is included in the condition list, the relatively common c.*97G>A (20210G>A) variant was excluded because it is associated with later-onset pro-thrombotic disease.^2^ |
| *FANCA* | Fanconi anemia, complementation group A | *FANCA-*related Fanconi anemia was excluded because bone marrow failure is rare in infancy, surveillance is not recommended to begin until later in childhood, and bone marrow harvest would not typically be undertaken in the first year of life.^3^ |
| *GBA1* | Gaucher disease, type I | *GBA1*-related Gaucher disease was excluded because the treatable form of this condition, type I Gaucher disease, is not typically actionable until after symptom onset, which most commonly occurs after the first year of life.^4^ |
| *LDLR* | Hypercholesterolemia, familial, 1 | LDLR-related Hypercholesterolemia was excluded because dietary therapy and statin therapy are typically not recommended in the first year of life.^5^ |

1. Valle, L., Katz, L. H., Latchford, A., Mur, P., Moreno, V., Frayling, I. M., Heald, B., Capellá, G., & InSiGHT Council (2023). Position statement of the International Society for Gastrointestinal Hereditary Tumours (InSiGHT) on *APC* I1307K and cancer risk. *Journal of medical genetics*, *60*(11), 1035–1043. <https://doi.org/10.1136/jmg-2022-108984>
2. Kujovich, J. L. (2006). Prothrombin Thrombophilia. In M. P. Adam (Eds.) et. al., *GeneReviews®*. University of Washington, Seattle.
3. Mehta, P. A., & Ebens, C. L. (2002). Fanconi Anemia. In M. P. Adam (Eds.) et. al., *GeneReviews*®. University of Washington, Seattle.
4. Grabowski G. A. (2008). Phenotype, diagnosis, and treatment of Gaucher's disease. *Lancet* (London, England), 372(9645), 1263–1271. <https://doi.org/10.1016/S0140-6736(08)61522-6>
5. Ison, H. E., Clarke, S. L., & Knowles, J. W. (2014). Familial Hypercholesterolemia. In M. P. Adam (Eds.) et. al., *GeneReviews*®. University of Washington, Seattle.

### Table S5. List of clinical experts and their specialties

| **Name** | **Specialty** |
| --- | --- |
| Amy E. Roberts | Cardiac disorders |
| Ada Hamosh | Cardiac disorders |
| Angela E. Lin | Cardiac disorders |
| Brittney Murray | Cardiac disorders |
| Deborah M. Mitchell | Endocrine disorders |
| Ingrid A. Holm | Endocrine disorders |
| Ari J. Wassner | Endocrine disorders |
| Raymond J. Kreienkamp | Endocrine disorders |
| Melanie Babinski | Endocrine disorders |
| Joseph A. Majzoub | Endocrine disorders |
| Christina Jacobsen | Endocrine disorders |
| Yee-Ming Chan | Endocrine disorders |
| Belinda S. Lennerz | Endocrine disorders |
| Jay R. Thiagarajah | Gastrointestinal disorders |
| Alanna Strong | Gastrointestinal disorders |
| Heidi L. Rehm | Hearing loss disorders |
| Elliott Kozin | Hearing loss disorders |
| Cynthia M. Powell | Hearing loss disorders |
| Vijay G. Sankaran | Hematologic disorders |
| Lisa R. Diller | Hereditary cancer predisposition syndromes |
| Sharon E. Plon | Hereditary cancer predisposition syndromes |
| David T. Miller | Hereditary cancer predisposition syndromes |
| Scott R. Plotkin | Hereditary cancer predisposition syndromes |
| Rebecca C. Hale | Inborn errors of immunity |
| Xiao P. Peng | Inborn errors of immunity |
| Jennifer M. Puck | Inborn errors of immunity |
| Charles J. Billington Jr | Inborn errors of immunity |
| Christine M. Seroogy | Inborn errors of immunity |
| Hannie Zomer-Bolanos | Inborn errors of immunity |
| Kathleen E. Sullivan | Inborn errors of immunity |
| Maleewan Kitcharoensakkul | Inborn errors of immunity |
| Manish J. Butte | Inborn errors of immunity |
| Rebecca A. Marsh | Inborn errors of immunity |
| Ottavia M. Delmonte | Inborn errors of immunity |
| Eyal Grunebaum | Inborn errors of immunity |
| Scott W. Canna | Inborn errors of immunity |
| Ivona Aksentijevich | Inborn errors of immunity |
| Raphaela Goldbach-Mansky | Inborn errors of immunity |
| Daniella M. Schwartz | Inborn errors of immunity |
| Craig D. Platt | Inborn errors of immunity |
| Samuel C.C. Chiang | Inborn errors of immunity |
| Matthew J. Kan | Inborn errors of immunity |
| Joshua D. Milner | Inborn errors of immunity |
| Roshini S. Abraham | Inborn errors of immunity |
| Shanmuganathan Chandrakasa | Inborn errors of immunity |
| Teresa K. Tarrant | Inborn errors of immunity |
| Rebecca D. Ganetzky | Inherited metabolic disorders |
| Jessica Gold | Inherited metabolic disorders |
| Lilian Downie | Medical genetics (overall list review) |
| Julie Fleischer | Medical genetics (overall list review) |
| Melissa A. Walker | Neurologic disorders |
| Anne Fulton | Ophthalmologic disorders |
| Emily M. Place | Ophthalmologic disorders |
| Alexander M. Holtz | Pulmonary disorders |
| Weizhen Tan | Renal disorders |
| Friedhelm Hildebrandt | Renal disorders |

### Table S6. Genes proposed but rejected for the BEACONS-NBS condition list for not meeting clinical inclusion criteria (inclusion criteria 1 and 2)

| **Gene** | **OMIM Phenotype** |
| --- | --- |
| *ABCA3* | Surfactant metabolism dysfunction, pulmonary, 3 |
| *ABCB11* | Cholestasis, progressive familial intrahepatic 2 |
| *ABCB4* | Cholestasis, progressive familial intrahepatic 3 |
| *ACAD8* | Isobutyryl-CoA dehydrogenase deficiency |
| *ACADS* | Acyl-CoA dehydrogenase, short-chain, deficiency of |
| *ACADSB* | 2-methylbutyrylglycinuria |
| *ACOX2* | Bile acid synthesis defect, congenital, 6 |
| *ACTG1* | Deafness, autosomal dominant 20/26 |
| *ADAR* | Aicardi-Goutieres syndrome 6 |
| *ALDH5A1* | Succinic semialdehyde dehydrogenase deficiency |
| *ALPL* | Hypophosphatasia, infantile |
| *AMACR* | Bile acid synthesis defect, congenital, 4 |
| *AMH* | Persistent Mullerian duct syndrome, type I |
| *AMHR2* | Persistent Mullerian duct syndrome, type II |
| *APOA5* | Hypertriglyceridemia, susceptibility to |
| *APOC2* | Hyperlipoproteinemia, type Ib |
| *ARNT2* | ?Webb-Dattani syndrome |
| *ARPC5* | Severe early onset systemic inflammation and autoimmunity |
| *ATP7B* | Wilson disease |
| *ATP8B1* | Cholestasis, progressive familial intrahepatic 1 |
| *AUH* | 3-methylglutaconic aciduria, type I |
| *B2M* | Immunodeficiency 43 |
| *BACH2* | Immunodeficiency 60 |
| *BRCA1* | Fanconi anemia, complementation group S |
| *BRCA2* | Fanconi anemia, complementation group D1 |
| *BRIP1* | Fanconi anemia, complementation group J |
| *C8G* | C8g deficiency |
| *CA2* | Osteopetrosis, autosomal recessive 3, with renal tubular acidosis |
| *CACNA1H* | Hyperaldosteronism, familial, type IV |
| *CACNA1S* | Malignant hyperthermia susceptibility 5 |
| *CCDC103* | Ciliary dyskinesia, primary, 17 |
| *CCDC39* | Ciliary dyskinesia, primary, 14 |
| *CCDC40* | Ciliary dyskinesia, primary, 15 |
| *CCDC65* | Ciliary dyskinesia, primary, 27 |
| *CCNO* | Ciliary dyskinesia, primary, 29 |
| *CCR2* | Interstitial lung disease and mycobacteriosis |
| *CD19* | Immunodeficiency, common variable, 3 |
| *CD247* | ?Immunodeficiency 25 |
| *CD274* | Autoimmune disease, multisystem, infantile-onset, 5 |
| *CD8A* | Immunodeficiency 116 |
| *CDCA8* | Congenital hypothyroidism |
| *CFAP298* | Ciliary dyskinesia, primary, 26 |
| *CFAP300* | Ciliary dyskinesia, primary, 38 |
| *CFB* | Complement factor B deficiency |
| *COL4A3* | Alport syndrome 3B, autosomal recessive |
| *COL4A4* | Alport syndrome 2, autosomal recessive |
| *COL4A5* | Alport syndrome 1, X-linked |
| *COPG1* | Combined immunodeficiency 38 |
| *COPZ1* | Neutropenia, severe congenital, 12, autosomal recessive |
| *COQ5* | ?Coenzyme Q10 deficiency, primary, 9 |
| *CP* | Aceruloplasminemia |
| *CR2* | Immunodeficiency, common variable, 7 |
| *CXCR2* | WHIM syndrome 2 |
| *DBF4* | Severe congenital neutropenia 12 |
| *DBR1* | Xerosis and growth failure with immune and pulmonary dysfunction syndrome;  Susceptibility to acute infection-induced encephalopathy type 11 |
| *DCLRE1B* | Dyskeratosis congenita, recessive 8 |
| *DDB2* | Xeroderma pigmentosum, group E, DDB-negative subtype |
| *DHCR7* | Smith-Lemli-Opitz syndrome |
| *DNAAF1* | Ciliary dyskinesia, primary, 13 |
| *DNAAF11* | Ciliary dyskinesia, primary, 19 |
| *DNAAF2* | Ciliary dyskinesia, primary, 10 |
| *DNAAF3* | Ciliary dyskinesia, primary, 2 |
| *DNAAF4* | Ciliary dyskinesia, primary, 25 |
| *DNAAF5* | Ciliary dyskinesia, primary, 18 |
| *DNAAF6* | Ciliary dyskinesia, primary, 36, X-linked |
| *DNAH11* | Ciliary dyskinesia, primary, 7, with or without situs inversus |
| *DNAH5* | Ciliary dyskinesia, primary, 3, with or without situs inversus |
| *DNAH9* | Ciliary dyskinesia, primary, 40 |
| *DNAI1* | Ciliary dyskinesia, primary, 1, with or without situs inversus |
| *DNAI2* | Ciliary dyskinesia, primary, 9, with or without situs inversus |
| *DNAJC19* | 3-methylglutaconic aciduria, type V |
| *DNAL1* | Ciliary dyskinesia, primary, 16 |
| *DNASE2* | Autoinflammatory-pancytopenia syndrome |
| *DPP9* | Hemophagocytic lymphohistiocytosis-like hyperinflammation;  Hatipoglu immunodeficiency syndrome |
| *DRC1* | Ciliary dyskinesia, primary, 21 |
| *DUOX2* | Thyroid dyshormonogenesis 6 |
| *EDA* | Ectodermal dysplasia 1, hypohidrotic, X-linked |
| *EDAR* | Ectodermal dysplasia 10B, hypohidrotic/hair/tooth type, autosomal recessive |
| *EDARADD* | Ectodermal dysplasia 11A, hypohidrotic/hair/tooth type, autosomal dominant |
| *EIF2B1* | Leukoencephalopathy with vanishing white matter 1, with or without ovarian failure |
| *EPO* | ?Diamond-Blackfan anemia-like |
| *ERCC2* | Xeroderma pigmentosum, group D |
| *ERCC3* | Xeroderma pigmentosum, group B |
| *ERCC4* | Xeroderma pigmentosum, group F |
| *ERCC5* | Xeroderma pigmentosum, group G |
| *ERCC6L2* | Bone marrow failure syndrome 2 |
| *FANCA* | Fanconi anemia, complementation group A |
| *FANCB* | Fanconi anemia, complementation group B |
| *FANCC* | Fanconi anemia, complementation group C |
| *FANCD2* | Fanconi anemia, complementation group D2 |
| *FANCE* | Fanconi anemia, complementation group E |
| *FANCF* | Fanconi anemia, complementation group F |
| *FANCG* | Fanconi anemia, complementation group G |
| *FANCI* | Fanconi anemia, complementation group I |
| *FANCL* | Fanconi anemia, complementation group L |
| *FARSA* | Rajab interstitial lung disease with brain calcifications 2 |
| *FCSK* | Congenital disorder of glycosylation with defective fucosylation 2 |
| *FOXI1* | Enlarged vestibular aqueduct |
| *FOXJ1* | Ciliary dyskinesia, primary, 43 |
| *FTCD* | Glutamate formiminotransferase deficiency |
| *GALE* | Galactose epimerase deficiency |
| *GAS8* | Ciliary dyskinesia, primary, 33 |
| *GATA4* | Atrioventricular septal defect 4 |
| *GBA1* | Gaucher disease, type I |
| *GCGR* | Mahvash disease |
| *GFUS** |  |
| *GHRHR* | Growth hormone deficiency, isolated, type IV |
| *GLA* | Fabry disease |
| *GLB1* | GM1-gangliosidosis, type I |
| *GM1* | GM1-gangliosidosis, type I |
| *GNE* | Nonaka myopathy |
| *GNMT* | Glycine N-methyltransferase deficiency |
| *GNPTAB* | Mucolipidosis II alpha/beta |
| *GNRH1* | Hypogonadotropic hypogonadism 12 with or without anosmia |
| *GPR101* | Pituitary adenoma 2, GH-secreting |
| *HCFC1* | Methylmalonic aciduria and homocysteinemia, cblX type |
| *HEATR3* | Diamond-Blackfan anemia 21 |
| *HELLS* | Immunodeficiency-centromeric instability-facial anomalies syndrome 4 |
| *HEXA* | Tay-Sachs disease |
| *HEXB* | Sandhoff disease, infantile, juvenile, and adult forms |
| *HGSNAT* | Mucopolysaccharidosis type IIIC (Sanfilippo C) |
| *HK1* | Hemolytic anemia due to hexokinase deficiency |
| *HMGCS2* | HMG-CoA synthase-2 deficiency |
| *HNF1A* | MODY, type III |
| *HNF1B* | Renal cysts and diabetes syndrome |
| *HNF4A* | MODY, type I |
| *HPS1* | Hermansky-Pudlak syndrome 1 |
| *HPS3* | Hermansky-Pudlak syndrome 3 |
| *HSD17B10* | HSD10 mitochondrial disease |
| *HSD17B4* | D-bifunctional protein deficiency |
| *HTRA2* | 3-methylglutaconic aciduria, type VIII |
| *HYDIN* | Ciliary dyskinesia, primary, 5 |
| *HYOU1* | Immunodeficiency 59 and hypoglycemia |
| *ICOS* | Immunodeficiency, common variable, 1 |
| *IFIH1* | Aicardi-Goutieres syndrome 7 |
| *IFNG* | Mendelian Susceptibility to mycobacterial disease 17 |
| *IKZF2* | Immunodysregulation, craniofacial anomalies, hearing impairment, athelia, and developmental delay |
| *IKZF3* | Immunodeficiency 84 |
| *IL10* | Il-10 deficiency |
| *IL18BP* | Hepatitis, fulminant viral, susceptibility to |
| *IL21R* | Immunodeficiency 56 |
| *IRF1* | Immunodeficiency 117 |
| *IRF7* | Immunodeficiency 39 |
| *IRF9* | Immunodeficiency 65 |
| *ITGAV* | Immune dysregulation, neurodevelopmental defects, and colitis |
| *ITPKB* | Combined immunodeficiency 35 |
| *KISS1R* | Hypogonadotropic hypogonadism 8 with or without anosmia |
| *KMT2B* | Intellectual developmental disorder, autosomal dominant 68 |
| *LAMTOR2* | Immunodeficiency due to defect in MAPBP-interacting protein |
| *LDLR* | Hypercholesterolemia, familial, 1 |
| *LDLRAP1* | Hypercholesterolemia, familial, 4 |
| *LIAS* | Hyperglycinemia, lactic acidosis, and seizures |
| *LMF1* | Lipase deficiency, combined |
| *LMNA* | Cardiomyopathy, dilated, 1A |
| *LPIN1* | Myoglobinuria, acute recurrent, autosomal recessive |
| *LRP4* | Cenani-Lenz syndactyly syndrome |
| *LRRC56* | Ciliary dyskinesia, primary, 39 |
| *LSM11* | Aicardi-Goutieres syndrome 8 |
| *LTBR* | Immunodeficiency with lymph node aplasia and hyposplenism |
| *LYN* | Systemic early-onset autoinflammation, vasculitis and hepatopathy |
| *MAD2L2* | Fanconi anemia, complementation group V |
| *MAGT1* | Immunodeficiency, X-linked, with magnesium defect, Epstein-Barr virus infection and neoplasia |
| *MAN2B2* | Combined immunodeficiency 37 |
| *MAP1LC3B2** |  |
| *MAP3K14* | Immunodeficiency 112 |
| *MAPK8* | JNK1 haploinsufficiency |
| *MAT1A* | Hypermethioninemia, persistent, autosomal dominant, due to methionine adenosyltransferase I/III deficiency |
| *MCIDAS* | Ciliary dyskinesia, primary, 42 |
| *MCM10* | Immunodeficiency 80 |
| *MCTS1* | Mendelian Susceptibility to Mycobacterial Disease 18 |
| *MRTFA* | Immunodeficiency 66 |
| *MT-TL1** |  |
| *MYO9A* | Myasthenic syndrome, congenital, 24, presynaptic |
| *NADK2* | 2,4-dienoyl-CoA reductase deficiency |
| *NAGLU* | Mucopolysaccharidosis type IIIB (Sanfilippo B) |
| *NEUROD1** |  |
| *NFATC1* | Immunodeficiency 111 with perturbed glycolysis |
| *NFKB1* | Immunodeficiency, common variable, 12 |
| *NFKB2* | Immunodeficiency, common variable, 10 |
| *NKX2-2* | Diabetes, permanent neonatal & neurological abnormalities |
| *NKX2-5* | Hypothyroidism, congenital nongoitrous, 5 |
| *NUDCD3* | Severe combined immunodeficiency 17 |
| *ODAD1* | Ciliary dyskinesia, primary, 20 |
| *ODAD2* | Ciliary dyskinesia, primary, 23 |
| *ODAD3* | Ciliary dyskinesia, primary, 30 |
| *ODAD4* | Ciliary dyskinesia, primary, 35 |
| *OPA3* | 3-methylglutaconic aciduria, type III |
| *PALB2* | Fanconi anemia, complementation group N |
| *PAX5* | Hypogammaglobulinemia, neuropathy and autism |
| *PAX6* | Cataract with late-onset corneal dystrophy; Aniridia |
| *PCSK9* | Hypercholesterolemia, familial, 3 |
| *PHKA1* | Muscle glycogenosis |
| *PIK3CG* | Immunodeficiency 97 |
| *PKD1* | Polycystic kidney disease 1 |
| *PKD2* | Polycystic kidney disease 2 |
| *PKHD1* | Polycystic kidney disease 4, with or without hepatic disease |
| *PLCG1* | ?Immune dysregulation, autoimmunity, and autoinflammation |
| *PLD4* | Systemic lupus erythematosus 18 |
| *PLOD2* | Bruck syndrome 2 |
| *PMM2* | Congenital disorder of glycosylation, type Ia |
| *PMVK* | Autoinflammation due to PMVK deficiency; Porokeratosis 1, multiple types |
| *POLD1* | Combined immunodeficiency 14A |
| *POLD3* | Immunodeficiency 122 |
| *POR* | Antley-Bixler syndrome with genital anomalies and disordered steroidogenesis |
| *PPM1K* | Maple syrup urine disease, mild variant |
| *PPOX* | Variegate porphyria, childhood-onset |
| *PPT1* | Ceroid lipofuscinosis, neuronal, 1 |
| *PSMA5** |  |
| *PSMB10* | Severe combined immunodeficiency 18;  Proteasome-associated autoinflammatory syndrome-5 |
| *PSMB4* | Proteasome-associated autoinflammatory syndrome 3 |
| *PSMB9* | Proteasome-associated autoinflammatory syndrome 3b;  Proteasome-associated autoinflammatory syndrome 6 |
| *PSMC10** |  |
| *PSMG2* | Proteasome-associated autoinflammatory syndrome 4 |
| *PTCRA* | Immunodeficiency 126 |
| *RFWD3* | Fanconi anemia, complementation group W |
| *RHBDF2* | iRHOM2 deficiency |
| *RHOG* | Hemophagocytic lymphohistiocytosis, familial, 8 |
| *RNASEH2A* | Aicardi-Goutieres syndrome 4 |
| *RNASEH2C* | Aicardi-Goutieres syndrome 3 |
| *RNU7-1* | Aicardi-Goutieres syndrome 9 |
| *RORC* | Immunodeficiency 42 |
| *RPL18* | ?Diamond-Blackfan anemia 18 |
| *RPL27* | ?Diamond-Blackfan anemia 16 |
| *RPL31* | Diamond-Blackfan anemia |
| *RPL35* | ?Diamond-Blackfan anemia 19 |
| *RPS15A* | Diamond-Blackfan anemia 20 |
| *RPS27* | ?Diamond-Blackfan anemia 17 |
| *RPS28* | Diamond Blackfan anemia 15 with mandibulofacial dysostosis |
| *RSPH1* | Ciliary dyskinesia, primary, 24 |
| *RSPH3* | Ciliary dyskinesia, primary, 32 |
| *RSPH4A* | Ciliary dyskinesia, primary, 11 |
| *RSPH9* | Ciliary dyskinesia, primary, 12 |
| *SAMHD1* | Aicardi-Goutieres syndrome 5 |
| *SCN2A* | Developmental and epileptic encephalopathy 11 |
| *SCN4A* | Myasthenic syndrome, congenital, 16 |
| *SCO2* | Mitochondrial complex IV deficiency, nuclear type 2 |
| *SEC61A1* | Immunodeficiency, common variable, 15 |
| *SECISBP2* | Thyroid hormone metabolism, abnormal, 1 |
| *SERAC1* | 3-methylglutaconic aciduria with deafness, encephalopathy, and Leigh-like syndrome |
| *SERPINA1* | Emphysema due to AAT deficiency |
| *SERPING1* | Angioedema, hereditary, 1 and 2 |
| *SFTPB* | Surfactant metabolism dysfunction, pulmonary, 1 |
| *SGSH* | Mucopolysaccharidosis type IIIA (Sanfilippo A) |
| *SHARPIN* | Autoinflammation with episodic fever and immune dysregulation |
| *SLC16A1* | Monocarboxylate transporter 1 deficiency |
| *SLC19A1* | Immunodeficiency 114, folate-responsive |
| *SLC25A32* | Exercise intolerance, riboflavin-responsive |
| *SLC26A7** |  |
| *SLC30A10* | Hypermanganesemia with dystonia 1 |
| *SLC3A1* | Cystinuria |
| *SLC4A4* | Renal tubular acidosis, proximal, with ocular abnormalities |
| *SLC52A1* | Riboflavin deficiency |
| *SLC7A9* | Cystinuria |
| *SLX4* | Fanconi anemia, complementation group P |
| *SMPD1* | Niemann-Pick disease |
| *SNAP25* | Developmental and epileptic encephalopathy 117 |
| *SNORA31* | Susceptibility to acute infection-induced encephalopathy type 10 |
| *SOCS1* | Autoinflammatory syndrome, familial, with or without immunodeficiency |
| *SOX3* | Panhypopituitarism, X-linked |
| *SPAG1* | Ciliary dyskinesia, primary, 28 |
| *SPPL2A* | Immunodeficiency 86, mycobacteriosis |
| *SRP68* | ?Neutropenia, severe congenital, 10, autosomal recessive |
| *STAT4* | Disabling pansclerotic morphea of childhood |
| *STXBP3* | Inflammatory Bowel Disease, Hearing Loss and Immune Dysregulation |
| *SYK* | Immunodeficiency 82 with systemic inflammation |
| *TACR3* | Hypogonadotropic hypogonadism 11 with or without anosmia |
| *TAPBP* | Bare lymphocyte syndrome, type I |
| *TCF3* | Agammaglobulinemia 8A, autosomal dominant;  Agammaglobulinemia 8B, autosomal recessive |
| *TERC* | Dyskeratosis congenita, autosomal dominant 1 |
| *TET2* | Immunodeficiency 75 |
| *TGFBR1* | Loeys-Dietz syndrome 1 |
| *TGFBR2* | Loeys-Dietz syndrome 2 |
| *THAP11* | ?Methylmalonic aciduria and homocystinuria, cblL type |
| *THRB* | Thyroid hormone resistance |
| *TIMM50* | 3-methylglutaconic aciduria, type IX |
| *TJP2* | Cholestasis, progressive familial intrahepatic 4 |
| *TLR3* | Immunodeficiency 83, susceptibility to viral infections |
| *TLR8* | Immunodeficiency 98 with autoinflammation |
| *TNFSF9* | Susceptibility to severe EBV infections 2 |
| *TRAC* | T-cell receptor-alpha/beta deficiency |
| *TRIM22* | Inflammatory bowel disease 32 |
| *TSPYL1* | Sudden infant death with dysgenesis of the testes syndrome |
| *TSR2* | ?Diamond-Blackfan anemia 14 with mandibulofacial dysostosis |
| *TTC7A* | Gastrointestinal defects and immunodeficiency syndrome |
| *UBE2T* | Fanconi anemia, complementation group T |
| *UNC93B1* | Early-onset TLR7-dependent autoimmunity;  Susceptibility to acute infection-induced encephalopathy type 1 |
| *USP53* | Cholestasis, progressive familial intrahepatic, 7, with or without hearing loss |
| *VKORC1* | Vitamin K-dependent clotting factors, combined deficiency of, 2 |
| *WFS1* | Wolfram syndrome 1 |
| *XPA* | Xeroderma pigmentosum, group A |
| *ZBTB7B** |  |
| *ZFPM2* | 46XY sex reversal 9 |
| *ZFYVE19* | Cholestasis, progressive familial intrahepatic, 9 |
| *ZMYND10* | Ciliary dyskinesia, primary, 22 |
| *ZNF143** |  |

**Condition does not have an OMIM entry*

### Table S7. Genes associated with secondary RUSP conditions which were excluded from Draft 2 of the BEACONS-NBS condition list

| **Gene** | **OMIM Phenotype** |
| --- | --- |
| *ACAD8* | Isobutyryl-CoA dehydrogenase deficiency |
| *ACADS* | Acyl-CoA dehydrogenase, short-chain, deficiency of |
| *ACADSB* | 2-methylbutyrylglycinuria |
| *AUH* | 3-methylglutaconic aciduria, type I |
| *DNAJC19* | 3-methylglutaconic aciduria, type V |
| *FTCD* | Glutamate formiminotransferase deficiency |
| *GALE* | Galactose epimerase deficiency |
| *GNMT* | Glycine N-methyltransferase deficiency |
| *HBA1* | Thalassemia, alpha- |
| *HBA2* | Thalassemia, alpha- |
| *HCFC1* | Methylmalonic aciduria and homocysteinemia, cblX type |
| *HSD17B10* | HSD10 mitochondrial disease |
| *HTRA2* | 3-methylglutaconic aciduria, type VIII |
| *MAT1A* | Hypermethioninemia, persistent, autosomal dominant, due to methionine adenosyltransferase I/III deficiency |
| *NADK2* | 2,4-dienoyl-CoA reductase deficiency |
| *OPA3* | 3-methylglutaconic aciduria, type III |
| *PPM1K* | Maple syrup urine disease, mild variant |
| *SERAC1* | 3-methylglutaconic aciduria with deafness, encephalopathy, and Leigh-like syndrome |
| *TIMM50* | 3-methylglutaconic aciduria, type IX |

### Table S8. Genes proposed but rejected from the BEACONS-NBS condition list due to concerns that detecting likely pathogenic and pathogenic variants would be technically challenging using whole genome sequencing (inclusion criterion 3)

| **Gene** | **OMIM phenotype** | **Reason for Exclusion** |
| --- | --- | --- |
| 11p15.5 region | Beckwith-Wiedemann syndrome | Methylation defects not detectable by sequencing |
| *ATAD3A* | Harel-Yoon syndrome | High paralog homology |
| *CD46* | Hemolytic uremic syndrome, atypical, susceptibility to, 2 | Limited predictive value for screening due to multifactorial inheritance |
| *CDKN1C* | Beckwith-Wiedemann syndrome | Insufficient coverage and methylation defects due to paternal imprinting not detectable by sequencing |
| *CFH* | Hemolytic uremic syndrome, atypical, 1;  Complement factor H deficiency | Limited predictive value for screening due to multifactorial inheritance |
| *CFI* | Hemolytic uremic syndrome, atypical, 3; Complement factor I deficiency | Limited predictive value for screening due to multifactorial inheritance |
| *CYP11B1* | Adrenal hyperplasia, congenital, due to 11-beta-hydroxylase deficiency | High paralog homology |
| *CYP11B2* | Hypoaldosteronism, congenital, due to CMO I deficiency | High paralog homology |
| *CYP21A2* | Hyperandrogenism, nonclassic type, due to 21-hydroxylase deficiency | Pseudogene interference |
| *CYP27B1* | Vitamin D-dependent rickets, type I | Common variant difficult to detect by WGS with no confirmation method from DBS |
| *FXN* | Friedreich ataxia | Repeat expansion not validated for detection by short read sequencing |
| *GFI1* | Neutropenia, severe congenital 2, autosomal dominant | Paternal imprinting requiring parental origin determination |
| *GH1* | Growth hormone deficiency, isolated, type IA | Limited ability to distinguish severe and milder phenotypes |
| *GNAS* | Pseudohypoparathyroidism Ia | Methylation defects due to paternal imprinting not detectable by sequencing |
| *HBA1* | Thalassemia, alpha- | High paralog homology and copy number variant detection limitations |
| *HBA2* | Thalassemia, alpha- | High paralog homology |
| *IGHM* | Agammaglobulinemia 1 | Reference genome mapping challenges |
| *IKBKG* | Immunodeficiency 33 | Pseudogene interference |
| *MNX1* | Currarino syndrome | GC rich region with poor coverage by WGS |
| *MT-RNR1** |  | Mitochondrial genome analysis not available |
| *NCF1* | Chronic granulomatous disease 1, autosomal recessive | Pseudogene interference |
| *PERCC1* | Diarrhea 11, malabsorptive, congenital | Reference genome mapping challenges and limited evidence supporting gene-disease relationship |
| *PHOX2B* | Central hypoventilation syndrome, congenital, 1, with or without Hirschsprung disease | Repeat expansion not validated for detection by short read sequencing |
| *PMS2* | Lynch syndrome 4 | Pseudogene interference and high paralog homology |
| *REST* | Wilms tumor 6, susceptibility to | Repetitive sequence region with alignment ambiguity |
| *RPS17* | Diamond-Blackfan anemia 4 | Copy number variant detection limitations |
| *SBDS* | Shwachman-Diamond syndrome 1 | High paralog homology |
| *SGLT1* | Glucose/galactose malabsorption | Duplicate of SLC5A1 |
| *SMAD4* | Juvenile polyposis/hereditary hemorrhagic telangiectasia syndrome | Pseudogene interference |
| *TREX1* | Aicardi-Goutieres syndrome 1 | Limited ability to distinguish pathogenic from benign variants |
| *TRIM28* | Wilms tumor 7 | Paternal imprinting requiring parental origin determination |

**Condition does not have an OMIM entry*

### Table S11. Conditions on the BEACONS-NBS condition list without a non-genetic suggestive sign, symptom or biomarker

| **Gene** | **Condition** |
| --- | --- |
| *ALDOB* | *ALDOB*-related hereditary fructose intolerance |
| *ALK* | *ALK*-related neuroblastic tumor susceptibility |
| *APC* | *APC*-related familial adenomatous polyposis |
| *BRAF* | *BRAF*-related RASopathy |
| *CTR9* | *CTR9*-related Wilms tumor |
| *DICER1* | *DICER1*-related pleuropulmonary blastoma familial tumor predisposition syndrome |
| *FGFR3* | *FGFR3*-related skeletal dysplasia |
| *FLAD1* | *FLAD1*-related flavin adenine dinucleotide synthetase deficiency |
| *G6PC1* | *G6PC1*-related glycogen storage disease 1 |
| *GPC3* | *GPC3*-related Simpson-Golabi-Behmel syndrome |
| *HRAS* | *HRAS*-related RASopathy |
| *IL1RN* | *IL1RN*-related interleukin 1 receptor antagonist deficiency |
| *JAG1* | *JAG1*-related Alagille syndrome |
| *KDM6A* | *KDM6A*-related Kabuki syndrome |
| *KMT2D* | *KMT2D*-related Kabuki syndrome |
| *KRAS* | *KRAS*-related RASopathy |
| *LEPR* | *LEPR*-related obesity |
| *LZTR1* | *LZTR1*-related RASopathy |
| *MAP2K1* | *MAP2K1*-related RASopathy |
| *MLH1* | *MLH1*-related constitutional mismatch repair deficiency |
| *MRAS* | *MRAS*-related RASopathy |
| *MSH2* | *MSH2*-related constitutional mismatch repair deficiency |
| *MSH6* | *MSH6*-related constitutional mismatch repair deficiency |
| *NF1* | *NF1*-related neurofibromatosis |
| *NOTCH2* | *NOTCH2*-related Alagille syndrome |
| *NRAS* | *NRAS*-related RASopathy |
| *OFD1* | *OFD1*-related ciliopathy |
| *PCK1* | *PCK1*-related cytosolic phosphoenolpyruvate carboxykinase deficiency |
| *PLPBP* | *PLPBP*-related vitamin B6-dependent epilepsy |
| *PTCH1* | *PTCH1*-related nevoid basal cell carcinoma syndrome |
| *PTPN11* | *PTPN11*-related Noonan syndrome with multiple lentigines |
| *PTPN11* | *PTPN11*-related RASopathy |
| *PYGL* | *PYGL*-related glycogen storage disease 6 |
| *RAF1* | *RAF1*-related RASopathy |
| *RB1* | *RB1*-related retinoblastoma |
| *RET* | *RET*-related multiple endocrine neoplasia 2B |
| *RIT1* | *RIT1*-related RASopathy |
| *RRAS2* | *RRAS2*-related RASopathy |
| *RYR1* | *RYR1*-related malignant hyperthermia |
| *SLC29A3* | *SLC29A3*-related histiocytosis-lymphadenopathy plus syndrome |
| *SLC5A6* | *SLC5A6*-related neurodegenerative disorder |
| *SMARCB1* | *SMARCB1*-related rhabdoid tumor predisposition syndrome and schwannomatosis spectrum |
| *SMN1* | *SMN1*-related spinal muscular atrophy |
| *SOS1* | *SOS1*-related RASopathy |
| *SOS2* | *SOS2*-related RASopathy |
| *SUFU* | *SUFU*-related nevoid basal cell carcinoma syndrome |
| *TP53* | *TP53*-related Li-Fraumeni syndrome |
| *TPK1* | *TPK1*-related thiamine pyrophosphokinase deficiency |
| *TSC1* | *TSC1*-related tuberous sclerosis complex |
| *TSC2* | *TSC2*-related tuberous sclerosis complex |
| *VHL* | *VHL*-related von Hippel-Lindau syndrome |
| *WT1* | *WT1*-related Wilms tumor |

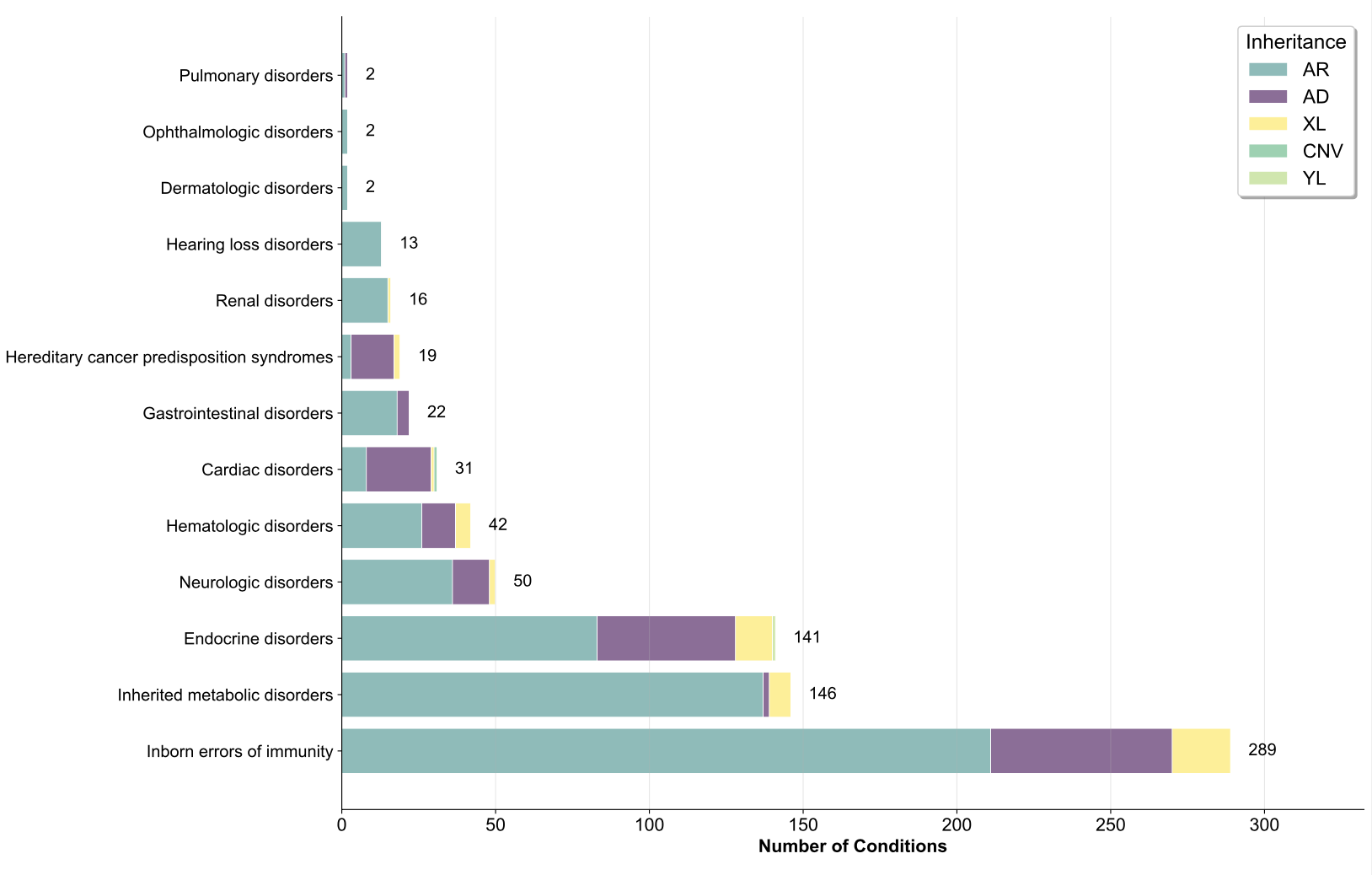

### Figure S1. Distribution of BEACONS-NBS conditions by clinical area and mode of inheritance. Bars represent the number of conditions within each clinical category. Colors indicate inheritance type.

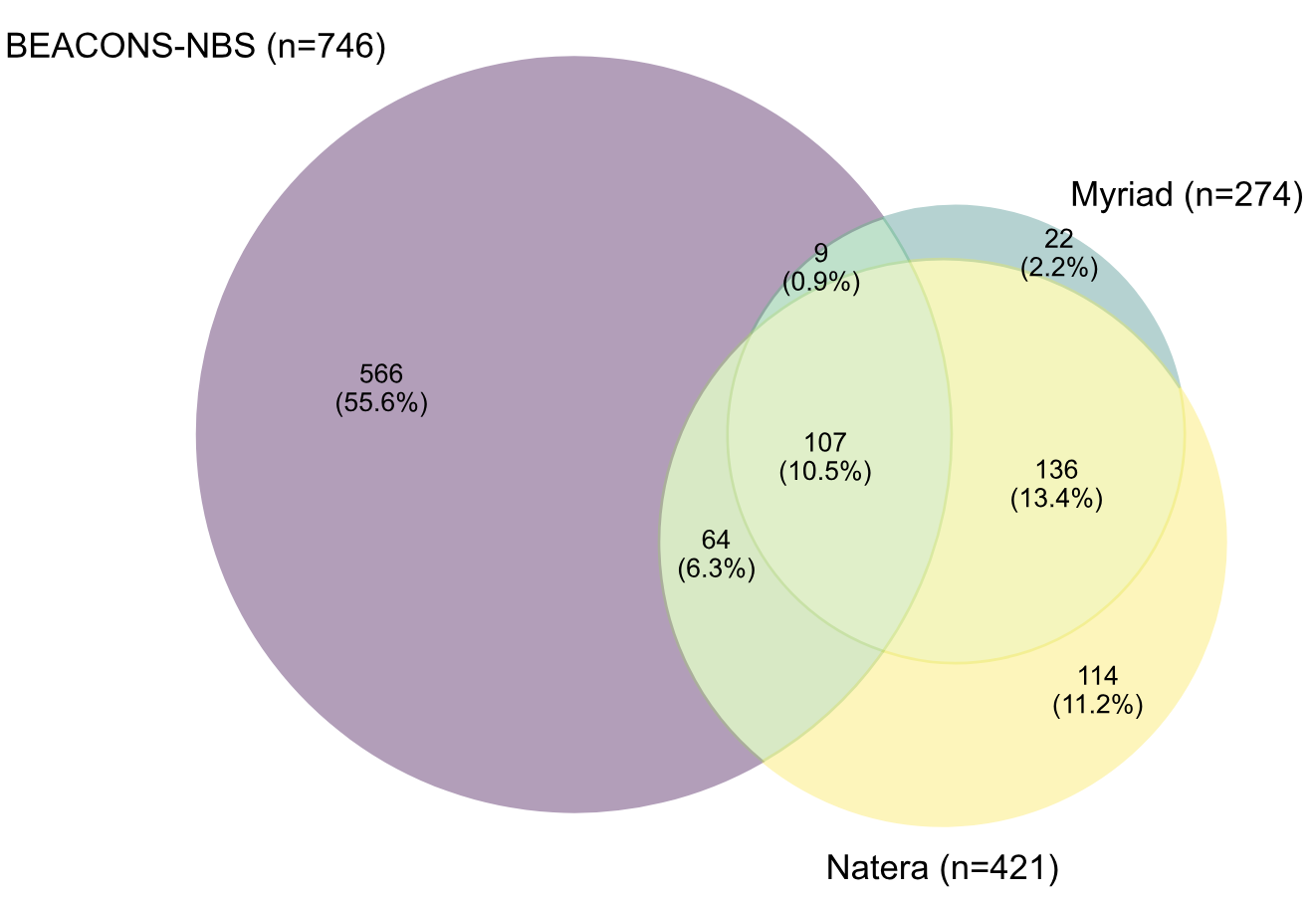

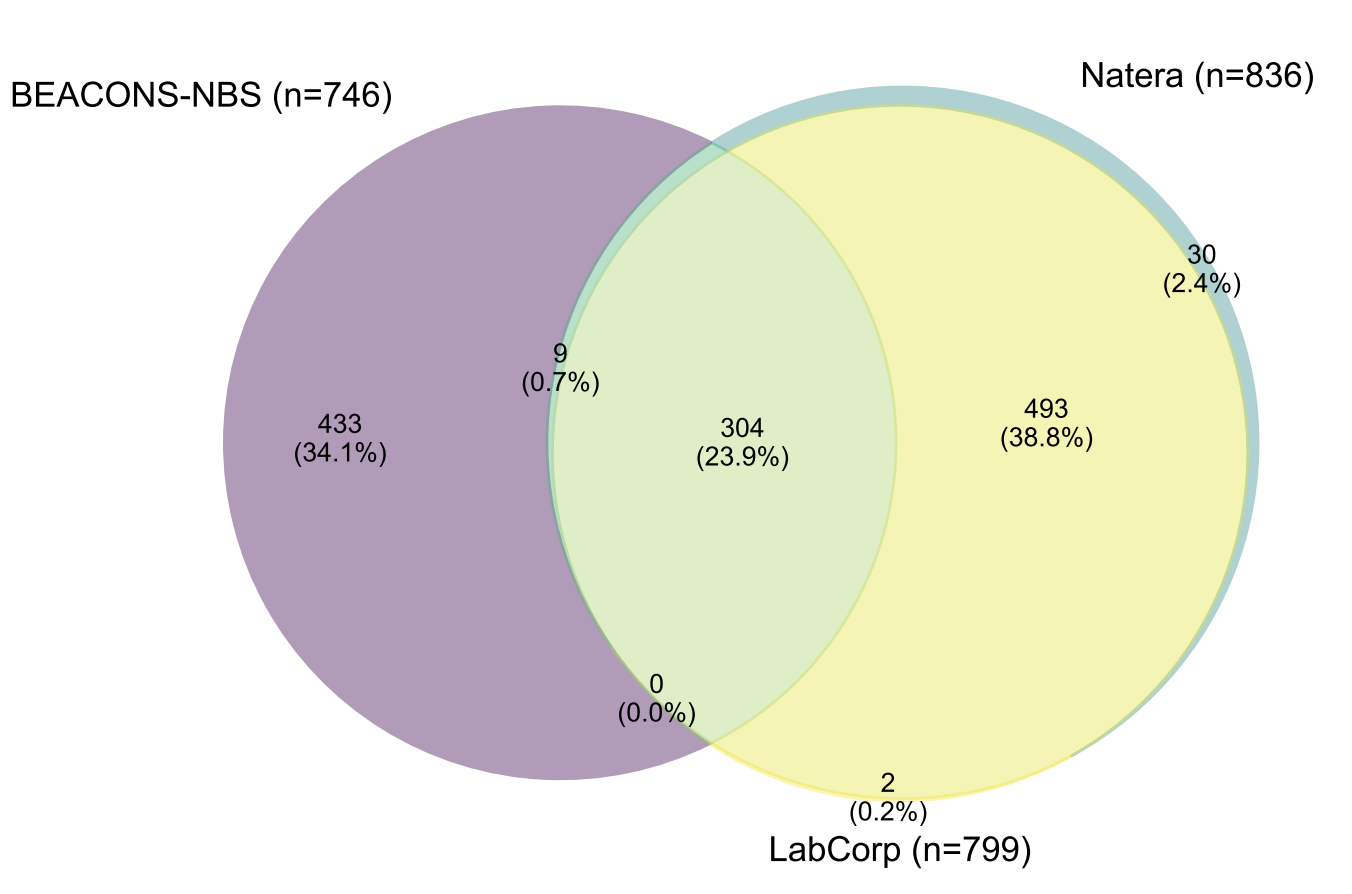

### Figure S2. Overlap of genes included in the BEACONS-NBS condition list and commercial carrier screening panels. A) Comparison of BEACONS-NBS with Myriad Foresight Carrier Screen and Natera Horizon 421 carrier screening panels. B) Comparison of BEACONS-NBS with Natera Horizon 835 and LabCorp 790 PLUS carrier screening panels.
